# Large-scale Proteomics Profiling of Peripheral Blood of DM1 patients identifies biomarkers for disease severity and functional capacity

**DOI:** 10.1101/2025.09.05.25335077

**Authors:** Daniël van As, Tine Claeys, Renee Salz, Delphi Van Haver, Sara Dufour, Amber van Deelen, Jolein Gloerich, Ralf Gabriels, Pieter Jan Volders, Vera Dobelmann, Andrea Gangfuss, Tobias Ruck, Genevieve Gourdon, Elise Duchesne, Cynthia Gagnon, Andreas Roos, Alain van Gool, Francis Impens, Lennart Martens, Hanns Lochmüller, Benedikt Schoser, Guillaume Bassez, Baziel G.M. van Engelen, Peter A.C. ’t Hoen, OPTIMISTIC consortium, ReCognitION consortium

## Abstract

**Background:** Myotonic Dystrophy Type 1 (DM1), the most common genetic neuromuscular disorder in adults, poses significant challenges for drug development due to its multisystem nature and high clinical variability in symptoms and disease progression. With a growing number of therapies entering clinical trials, this study addresses the urgent need for biomarkers that can serve as surrogate endpoints.

**Methods:** We profiled 437 serum samples from adult DM1 patients collected at two timepoints of the OPTIMISTIC trial using bottom-up mass spectrometry with data-independent acquisition. Associations between protein expression, the disease-causing CTG-repeat and 25 clinical outcome measures were studied using linear mixed-effect models. All key study findings were validated in an independent cohort of 69 DM1 patients and 10 healthy controls.

**Results:** Of the 259 identified proteins, 161 showed significant associations with the CTG-repeat length (FDR < 5%). Hypogammaglobulinemia was confirmed and shown to be worse in severely affected patients. A strong proteomic signature was associated with clinical measures of functional capacity, with the 6-Minute Walk Test showing the strongest signal (70 associations, FDR < 5%). These novel associations reveal a compelling link between chronic inflammation and reduced functional capacity. A machine learning algorithm identified a minimal set of 13 proteins robustly reflecting both the underlying genetic defect and functional capacity.

**Conclusions:** DM1 induces a broad disease fingerprint in the serum proteome, predominantly affecting proteins of the immune system. A carefully selected panel of proteins showed the greatest potential to meet the statistical criteria required for surrogate endpoints in clinical trials.

## Background

Myotonic Dystrophy type 1 (DM1) is a progressive, multisystem disease with an estimated prevalence of 1 in 8,000, with some studies reporting a locally much higher prevalence such as 1 in 550 in the Saguenay-Lac-Saint-Jean area in Northeastern Quebec and 1 in 2,100 for the state of New York^1,2^. Various organ systems are involved in the disease, causing symptoms such as muscular weakness, myotonia, severe fatigue, apathy, cataracts, diabetes and various respiratory, cardiac and gastrointestinal problems. DM1 is caused by a trinucleotide expansion of more than 50 CTG repeats in the DM1 protein kinase (*DMPK*) gene^3–5^. While often serving as a proxy for disease severity, the magnitude of the (progenitor) CTG repeat expansion has only a moderate negative correlation with the age of disease onset and a moderate positive association with the severity of the clinical phenotype^6^. Furthermore, it has been shown that interruptions of the CTG-repeat by variant repeats, such as CCG or CGG, are associated with a milder clinical phenotype^7^.

The largest randomized controlled clinical trial in DM1, OPTIMISTIC, has investigated the effect of personalized Cognitive Behavioural Therapy (CBT), with the optional addition of graded exercise therapy, in a cohort of more than 250 genetically characterized DM1 patients. While it has been shown that CBT can, on average, slightly but significantly improve the capacity for activity and participation of DM1 patients, the magnitude of the improvements and the improved disease aspects were highly heterogenous^8,9^. This result highlights an important limitation in current DM1 clinical trial research. The heterogeneity of the patient population asks for more targeted clinical trials of patient subpopulations with shared disease characteristics. However, patient subpopulations are currently not well-defined, and subgroup analyses in a classical statistical framework may suffer from reduced statistical power.

In novel clinical trial designs, biomarkers are a valuable addition to the evaluation of therapy effects. Given the shared molecular dysregulations induced by the CTG expansion, one may expect more homogenous molecular responses to an intervention in comparison to clinical responses. Furthermore, these molecular changes may be more sensitive to change, preceding clinical benefits such as delayed disease progression. Since an ideal biomarker can be efficiently obtained with a low burden for patients, analyses of (peripheral) blood have sparked great interest in biomarker research. For various neurological and psychiatric disorders, including Duchenne Muscular Dystrophy, Huntington’s disease, major depressive disorder and DM1, disease-relevant findings have been reported in blood^10–13^. This finding aligns with our earlier transcriptomic research of the OPTIMISTIC cohort, where we identified significant associations between the CTG repeat and 608 genes expressed in peripheral blood, of which 97 returned to more normal expression levels in patients who clinically improved^14^.

Here, we expanded on this work by performing proteomic analysis of 437 serum samples collected at two timepoints of the OPTMISTIC trial. The most considerable differences in protein expression between patients were linked to the size of the CTG-repeat and confirmed the known hypogammaglobulinemia in DM1^15–17^. A strong proteomic fingerprint was also associated with multiple measures of functional capacity, showing primarily an increase in components of the complement system in patients with reduced functional performance. Rather than relying on individual associations with disease pathology or physical activity, we demonstrate that a set of 13 proteins collectively has the strongest potential to meet the criteria for surrogate endpoints in clinical trials. All key study findings, including the set of 13 proteins, were independently validated in an extensive cohort of 69 patients with DM1 and 10 healthy controls.

## Methods

### Sample sources

#### OPTIMISTIC clinical trial samples

All primary analyses were based on the samples and metadata that were collected during the OPTIMISTIC clinical trial (NCT02118779)^8,18^. In this European multi-centre clinical trial, 255 genetically confirmed adult DM1 patients across four European countries were enrolled. Patients were randomized either into the intervention arm (n=128), consisting of personalized cognitive behavioural therapy (CBT) with optionally the addition of graded exercise therapy (GET, n=32), or the control arm (n=127), consisting of standard of care. In both study arms, a rich phenotypic characterisation was obtained by assessing more than 25 outcome measures. We grouped these outcome measures into 6 disease domains (Functional capacity; Fatigue; Social Functioning, Behaviour and Impact on Caregivers; Cognitive Function, Depression, Illness Coping; Disease Impact, Severity and Pain; Apathy) and listed them together with their abbreviations in Table 1. Details of the individual outcome measures, including scoring ranges, have been published elsewhere^8,9^. Additionally, whole blood was collected at multiple timepoints for biomarker analyses (10 ml for DNA analysis, 10ml for RNA analysis and 10ml serum). Serum was isolated from serum-specific BD Vacutainer tubes (Ref 368815) and centrally stored at -80°C at the John Walton Muscular Dystrophy Research Centre Biobank^19^. Serum samples obtained at the start of the trial (n= 252) and at the primary trial endpoint after 10 months (n= 211) were sent to the Department of Biomolecular Medicine at Ghent University, Belgium, for mass-spectrometry based proteome analyses.

**Table 1:**
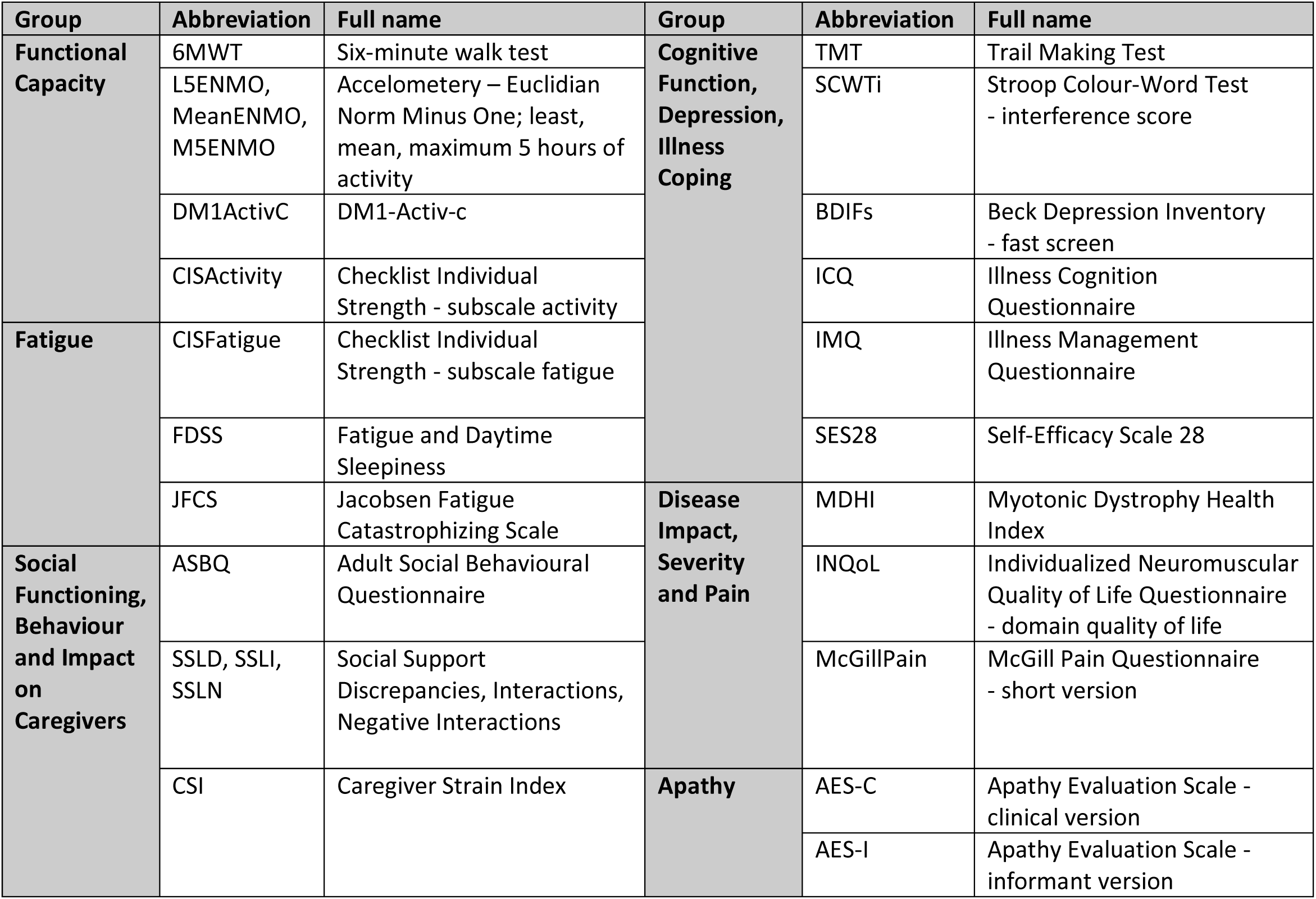
Overview of OPTIMISTIC outcome measures.

#### Canadian and German cohort samples

External validation of the study findings was implemented on a cohort of Canadian (n= 56) and German (n= 13) DM1 patients. DM1 patients from the Canadian cohort were recruited between 2011-2013 as part of a larger longitudinal study, all of whom having participated in the first phase of the study between 2002 and 2004. Inclusion criteria were to have the late-onset, adult or juvenile phenotype of DM1 confirmed by genetic analysis and to be aged 18 years or older. CTG repeat length was determined using the same methodology as that used in previous studies^20,21^. All clinical assessments were performed by the same physiotherapist. The 6-Minute Walk Test (6MWT) was performed, where the maximum distance walked along a 30-meter corridor over a 6-minute period was measured (in meters). Grip strength was assessed using a Jamar digital dynamometer (Asimow Engineering Co., Los Angeles, CA). The mean of three trials was used for analyses. DM1 patients from the German cohort were recruited between 2019 and 2024 in the Department of Pediatric Neurology of the University Hospital Essen (University Duisburg-Essen, Germany). Inclusion criteria were clinical and genetically confirmed DM1. This cohort included paediatric cases (juvenile DM1 patients) as well as diseased parents (adult DM1 patients). Moreover, serum samples of 10 healthy donors were collected from the same German site (clinically unaffected family members and unrelated individuals).

#### Mass-spectrometry based proteome analysis of the OPTIMISTIC cohort

Undepleted serum samples contain proteins in a large dynamic range, making the identification of low-abundant proteins challenging. To maximize the number of quantified serum proteins, a state-of-the-art data independent acquisition (DIA) mass-spectrometry-based workflow for clinical proteomics was applied at Ghent University. The recorded spectra were analysed using the DIA-NN software employing the built-in spectral library prediction^22^. Since peptides are quantified in MS-based proteomics, proteins must be inferred by matching these peptides to known sequences. When peptides could not be uniquely assigned to a single protein, a protein group is reported instead. This group encompasses all protein matches, thereby preventing the overestimation of protein identifications. Out of the 463 samples that were processed in this workflow, 11 samples were lost due to corrupt or incorrectly recorded MS datafiles (n=452 samples remaining). For a detailed workflow of the proteome quantification analysis, including sample preparation, LC-MS/MS analysis and proteomics data analysis, please refer to the ‘Mass spectrometry-based protein quantification of the OPTIMISTIC samples’ section in the appendix.

#### Proteomic analysis of the Canadian and German cohort

The proteomes of the Canadian (n=56) and German (n=23, including n=10 healthy controls) sera samples were assessed using a (DIA) mass-spectrometry-based workflow for clinical proteomics at the Radboud University Medical Center in Nijmegen. Recorded spectra were then analysed in real-time using the Proteoscape-implemented version of DIA-NN^23^. Similar to the DIA-NN based workflow applied to the OPTIMISTIC data, peptides that could not be uniquely matched to a specific protein were instead associated with a protein group. For a detailed workflow of the proteome quantification analysis, including sample preparation, LC-MS/MS analysis and proteomics data analysis, please refer to the ‘Mass spectrometry-based protein quantification of the Canadian and German cohort samples’ section in the appendix.

#### ELISA-based validation of ITIH3 serum levels

Additionally, for the Canadian sera samples (n=56), ELISA-based ITIH3 levels were measured in using the “Human inter-alpha-trypsin inhibitor heavy chain H3 (ITIH3) ELISA kit” (CSB-EL011896HU, Cusabio). The assay was performed at the Department of Neurology of the University Hospital Düsseldorf (Germany) according to the manufacturer’s protocol. In brief, samples and standards were added to the precoated plate, followed by addition of horseradish peroxidase conjugate. The plate was then incubated for 30 min at 37°C. The wells were washed five times, and TMB substrate was added. The plate was then incubated for 20 min at 37°C. Subsequently, the stop solution was added to end the reaction, and the optical density was measured with a microplate reader (Tecan) at 450 nm. Samples were used in a dilution of 1:500 and measured in duplicate.

### Ethics approval and consent to participate

#### OPTIMISTIC clinical trial samples

The OPTIMISTIC clinical trial (NCT02118779) was conducted in accordance with the Declaration of Helsinki and approved by the medical-ethical scientific committee for human research at each of the four participating clinical centres. Prior to the trial, all enrolled patients provided written informed consent, which included the usage of the pseudonymized blood samples for the research purposes of this study. Ethical approval for mass spectrometry-based proteomics profiling of the serum samples was obtained from the Ethics Committee of Ghent University Hospital (B670201940027). For more specific methodological details of the clinical trial, including trial protocols and an overview of all (patient-reported) outcome measures, please refer to the published trial protocol and the main study publication^8,18^.

#### Canadian cohort samples

The study was conducted at the Saguenay Neuromuscular Clinic and was approved by the Ethics Review Board of the Centre Intégré Universitaire de Santé et Services Sociaux du Saguenay–Lac-St-Jean (Chicoutimi, Québec, Canada; #2010-046). Written informed consent was obtained from all participants including biomarkers studies.

#### German cohort samples

All patients and/or caregivers, as well as healthy donors, gave written consent to donate blood samples for research-driven biomarker studies. The local ethical committee approved biomarker studies on neuromuscular patients and controls (19-9011-BO).

### Availability of data and materials

#### OPTMISTIC clinical trial

Researchers interested in accessing the phenotype data from the OPTIMISTIC study are invited to contact K. Mul and complete a data access agreement. All requests will be reviewed by a panel comprising from each of the four participating clinical sites, with K. Mul serving as chair^8^. The mass spectrometry proteomics data of the OPTIMISITIC samples have been deposited to the ProteomeXchange Consortium via the PRIDE partner repository with the dataset identifier PXD067476^24^.

#### External cohorts

The phenotype data used and/or analysed from the Canadian cohort are available from the corresponding author upon reasonable request following the proper evaluation of the research protocol by the Ethics Review Board of the Centre intégré universitaire de santé et de services sociaux du Saguenay–Lac-St-Jean (Saguenay, Québec, Canada). The phenotype data used from the German cohort, including the ELISA-based quantification data of ITIH3, are stored at the Department of Neurology of the University Hospital Düsseldorf (Heinrich Heine University) and are available upon request to.The mass spectrometry proteomics data of both the Canadian and German cohort have been deposited to the ProteomeXchange Consortium via the PRIDE partner repository with the dataset identifier PXD060035^24^.

#### Results and Code availability

For both studies, the full list of significant protein group associations with the CTG-repeat and 6MWT scores, as well as the table containing all Variable Inclusion Probabilities, will be made available on GitHub after publication. Additionally, all R scripts used in this work are available via https://github.com/cmbi/DM1_ReCognitION_Proteomics

### Data analysis

All analyses were implemented with the statistical programming language R within the RStudio integrated development environment^25,26^. All figures have been generated using the ggplot2 package and arranged using the cowplot package^27,28^. All selected candidate protein group biomarkers have been associated with gene names using UniProt^29^. Where appropriate, data presented in tabular format has been rounded to 4 digits.

#### Outlier handling of the phenotype data

For all outcome measures, we calculated the interquartile range (IQR) and considered measurements with Q1 – 3 * IQR and Q3 + 3 * IQR as potential outliers. Subsequently, these potential outliers were visually inspected to confirm the likelihood of an unreasonable or faulty measurement that could potentially skew the results of the statistical frameworks. For the 25 outcome measures of the OPTIMISTIC data, a total of 10 outliers were set to ‘NA’ (0.09% of the screened datapoints). For the Canadian cohort, a total of two outliers were set to ‘NA’ concerning the grip strength (left and right) of one unusually strong patient.

#### Pre-processing of raw peptide and protein group intensities

For the OPTIMISTIC study, pre-processing of both the peptide and protein group data has been implemented; however, the Canadian cohort validation study focused only on the protein group data. For the OPTIMISTIC peptide samples, all identified peptides with different post-translational modifications (PTMs) were summed up and total intensity values of less than 10,000 per peptide were changed to ‘NA’. To identify samples with low/unreliable peptide detection rates, total peptide abundance per sample was calculated by summing up all detected peptides. Ten samples with total intensities lower than Q1 – 3 * IQR were removed. Five of these samples were from the same row of a well plate, indicating a possible dilution error. Since the OPTIMSTIC protein group data were inferred based on the peptide data, these samples were also removed from the protein group dataset (n=442 samples remaining).

For the protein group data of both studies, intensities of less than 10,000 per protein group were set to ‘NA’. Technical replicates, which were only present in the external validation dataset, were averaged. For the OPTIMISTIC dataset, protein groups and associated peptides linked to the expected contaminants trypsin and bovine albumin were excluded. In contrast, for the Canadian cohort, the iRT-Kit protein group was removed. Next, for all the datasets, intensity values were log2 transformed and all samples were subsequently scaled using the ‘equalMedianNormalization’ function of the R package DEqMS (independently per dataset)^30^.

Peptides or protein group measurements with intensities lower than Q1 – 3 * IQR for that particular peptide or protein group across samples were removed. For the OPTIMISTIC samples, 2916 peptide measurements (0.29% of screened datapoints) and 141 protein group measurements (0.13% of screened datapoints) were set to ‘NA’ because of substantially different abundances across samples. Likewise, for the external validation dataset, 29 protein groups were set to ‘NA’ (0.22% of screened datapoints). During the last filter step, peptides or protein groups identified in fewer than 100 (OPTIMISTIC data, 23%) or 20 (external validation data, 36%) samples were removed because they are neither useful as biomarkers nor for robust statistical inference. In doing so, 25 peptides (n=2670 remaining) and one protein group (n=259 remaining) of the OPTIMISTIC data and 223 protein groups (n=259 remaining) of the external validation data were removed.

As a quality control procedure, mean-variance plots were generated for the protein group datasets. For the 189 paired OPTIMISTIC samples (from the same patient, at two timepoints), the Pearson correlation was calculated for each protein group between the baseline and the matched 10-month sample. This was compared with the average Pearson correlation of each protein group between the baseline and all other 10-month samples. For five baseline samples of the OPTIMISTIC datasets, no phenotype data were present, as these patients were ultimately not enrolled in the clinical trial. Accordingly, these peptide and protein group samples were removed, resulting in a final sample size of n=437.

An additional dataset was analysed based on the combined set of the Canadian (n=56) and German (n=23, including 10 healthy controls) samples. The pre-processing of this combined cohort was similar to the protein group datasets above, except that no intensity minimum of 10,000 was set and no outliers were removed. This dataset of 79 samples (comprising both Canadian and German cohorts) was exclusively used for the comparison of the top identified biomarkers in DM1 patients (n=69) versus healthy controls (n=10).

#### Principal component analyses

Principal component analyses (PCA) were implemented independently for the full peptide and protein group datasets of the OPTIMISTIC study, combining samples from both timepoints. Because PCA requires complete datasets, samples within the 25^th^ quantile for the number of identified peptides or protein groups were filtered out (respectively 327 and 315 samples remaining for the peptide and protein group data). Subsequently, both the peptide and protein group datasets were filtered for molecules present in all filtered samples (901 peptides and 173 protein groups remaining). The PCA was implemented using the R function ‘prcomp’ with the arguments ‘center’ and ‘scale’ set to TRUE. To study the amount of variance of each principal component that can be attributed to different phenotype measures, mixed effect models were fitted for the first 10 principal components using the ‘lmer’ function of the lme4 package with default settings^31^ [1].

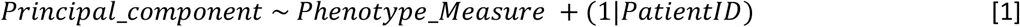

Marginal R-squared values (R-squared values attributable to the fixed effects) were obtained using the ‘r.squaredGLMM’ function of the MuMIn package^32^ and associations were visualized using the ggcorrplot package^33^.

#### Statistical Analyses

For the OPTIMISTIC protein group data, four different statistical mixed effect models were implemented for each protein group to respectively study the association with the CTG repeat [2], associations with different outcome measures [3], the effect of the cognitive behavioural therapy (CBT) [4] and the effect of the graded exercise therapy (GET) [5]. To account for proteomic differences attributable to biological sex, the covariate ‘Sex’ was included as fixed effect in all models. To account for the dependency of measures from the same patients and well plates, the variables ‘PatientID’ and ‘Plate’ were included as random effects in all models. In model [2], the same CTG repeat length was used at both timepoints, as the CTG-repeat size was not re-estimated after the primary trial endpoint. In models [2] and [3], the variable ‘Timepoint’ (before/after the trial) was included as a fixed effect to account for possible temporal changes that occurred during the trial. All models were generated using the ‘lmer’ function of the lme4 package with default settings^31^. P-values associated with the variables of interest (bold in the formulae) were generated with the lmerTest package and multiple testing corrected based on the Benjamini & Hochberg procedure using the R base ‘p.adjust’ function with method=‘fdr’^34^. Additionally, for all associations studied with models [2] and [3], Pearson correlations were calculated between the variable of interest and the protein group using the R base ‘p.cor’ function with use= ‘pairwise.complete.obs’ and method= ‘pearson’. Fits with convergence problems were automatically removed. As a measure of statistical robustness, the number of reported significant association with the CTG-repeat [2] and the 6MWT [3] are also reported after randomization of these two measures across the patients. Due to the unequal distribution of samples across well plates with respect to study timepoint, models [4] and [5] were additionally implemented without the random plate effect as a supplemental analysis to assess the potential confounding influence of plate-specific variation on treatment effects.

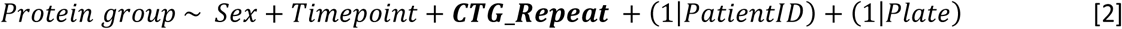

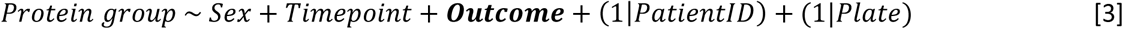

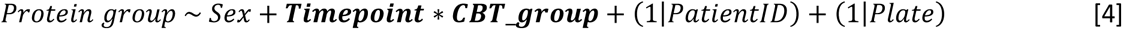

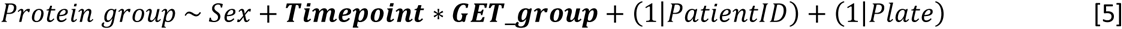

For the external validation study (Canadian cohort, n= 56), three linear models were fit to study the association of physical outcome measures (6MWT, grip strength left and right hand) with the ELISA-based ITIH3 levels [6]. Furthermore, for each MS-based identified protein group a linear model was fit to study the association with the CTG repeat [7] and 6MWT score [8]. All models included the covariate ‘Sex’ to account for possible differences attributable to biological sex. Plots were generated to illustrate the associations between log2 ITIH3 (and other candidate biomarkers) and the clinical scores. Additionally, for the associations between the variables of interest and the protein groups in models [7] and [8], Pearson correlations were calculated using the R base ‘p.cor’ function with use= ‘pairwise.complete.obs’ and method= ‘pearson’. Although adjusted p-values (Benjamini Hochberg procedure) for the results from models 7 and 8 were calculated, the validation analysis reported in this study focuses on the nominal p-values of the already multiple testing corrected top hits from the OPTIMISTIC study-based results.

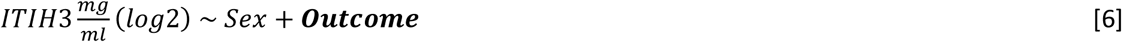

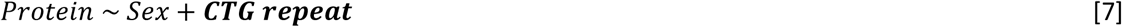

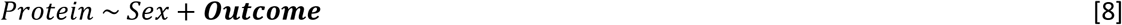

To externally validate the statistical model hits, we compared the FDR-adjusted significant protein group associations with the CTG repeat length and 6MWT from the OPTIMISTIC cohort [2,3] with the nominally significant associations observed in the external Canadian validation cohort [7,8]. However, direct comparison of protein groups between the OPTIMISTIC and Canadian cohorts was not feasible due to slight differences in their protein compositions. To address this, all protein groups were decomposed into individual proteins. Next, a non-redundant list of overlapping significant proteins was generated. For each protein, the most significantly associated protein group in the OPTIMISTIC cohort was identified. An optimal matching protein group from the Canadian cohort was then selected based on maximal overlap in constituent proteins. In cases where multiple external protein groups exhibited the same degree of overlap with a given OPTIMISTIC protein group, the group with the fewest total proteins was selected. If redundancy persisted, the most statistically significant external protein group was chosen. Finally, only matches with Pearson correlations in the same direction were retained.

For the models [2, 3, 4, 5], Volcano plots were generated, where for models [2, 3] additional colour coding was used to illustrate if the results were also nominally significant in the respective models [7, 8]. For models [2, 3], example plots of the strongest associations that were also externally validated were generated. To visualize the possible impact of CTG repeat interruptions (repeat variants) on the protein group-CTG repeat associations, the identified top immunoglobulin and complement-associated protein groups identified in model [2] were visualized with the color-coding reflecting whether a repeat interruption was present. For the models [4, 5] example plots were generated for the significant results (adjusted p < 0.05).

For the most promising biomarkers, differences in expression (healthy vs DM1) plots were generated based on the unfiltered combined Canadian (n=56) and German samples (13 DM1 patients, 10 healthy controls). Significance labels were added using the ‘stat_compare_means function (method= “t.test”, paired= “F”) of the ggpubr package^35^.

#### Bootstrap enhanced multivariate Elastic-Net regression

To identify a minimum subset of protein groups linked with both the disease pathology and the clinical phenotype, a multivariate Elastic-Net regression framework has been implemented. First, the 47 protein groups that were significantly associated with both the CTG-repeat expansion and the 6MWT, as determined by the statistical analyses of the OPTIMISTIC cohort, were selected as candidate predictors. To validate the results of this statistical framework, the 47 protein groups were reduced to 42 protein groups that were also unambiguously identified in the Canadian cohort. Model training was exclusively done using the OPTIMISTIC (baseline) dataset. The results were internally validated using the 10-month data (internal testing) and externally validated with the completely independent Canadian cohort.

Since the Elastic-Net framework necessitates a complete dataset, 10 imputed OPTIMISTIC protein group datasets were generated using Multiple Imputation Using Chained Equations (MICE)^36,37^. For the data imputation, the whole protein group dataset, spanning both timepoints, was used to allow for the most accurate results. Protein groups missing in more than 20% of the samples were excluded. For the remaining protein groups(n=239), each missing value was imputed based on the 100 protein groups with the highest associated absolute Pearson correlation, with 50 iterations per dataset.

The multivariate variable selection algorithm was then implemented on the imputed baseline datasets from the OPTIMISTIC study (model training) for cases with no missing CTG-repeat of 6MWT information (n=231). Given the large distribution differences of the dependent variables (CTG-repeat, 6MWT) between the OPTIMISTIC and external validation study, as well as the multivariate Gaussian framework of the Elastic-Net regression, the dependent variables were scaled using the ‘boxcox’ function of the R MASS package with automatically derived optimal lambda values^38^. Next, for each imputed dataset, 5000 bootstrap distributions were generated using the ‘boot’ function of the package boot^39^. Subsequently, for each bootstrap distribution, the ‘cv.glmnet’ function of the glmnet package, using the parameters type.measure=“mse”, family = “mgaussian”, alpha=0.5, nfolds=10, standardize=TRUE and standardize.response=TRUE, was implemented^40^. Coefficients were selected based on the “lambda.1se” setting, allowing for a minimum number of predictors that still perform within one standard error of the model with the best performance. For each imputed dataset, it was calculated how frequently a protein group has been selected across the 5000 bootstrap distributions, which is referred to as Variable Inclusion Probability (VIP)^9,41,42^. Finally, the VIPs across all imputed datasets were averaged, yielding a single average VIP for each protein group. A set of candidate predictors could then be obtained by selecting all protein groups that have a certain number of average VIPs.

The predictive power of a combined set of protein groups was systematically assessed using different average VIP thresholds (90%, 80%, 70% and 60%). For this, a regular multivariate linear regression framework was fitted on the unimputed OPTIMISTIC baseline data, where the dependent variables (CTG-repeat and 6MWT) were predicted based on the combined set of protein groups (baseline model). The baseline model was then used to predict the baseline values, the 10 month OPTIMISTIC values (unseen, internal validation), and the values from the Canadian cohort (unseen, external validation). In line with the variable selection algorithm, for all three datasets (OPTIMISTIC baseline and 10 months, external validation) the dependent variables were independently scaled using the ‘boxcox’ function of the R MASS package with automatically derived optimal lambda values and subsequently together with the independent variables (candidate protein groups) z-score transformed using the R-base ‘scale’ function with the default parameters center=True and scale=True^38^. (Adjusted) R-squared values of the baseline model are reported, as well as the Root Mean Square Error (RMSE) values for the predictions of the OPTIMISTIC baseline, OPTIMISTIC 10-month and Canadian cohorts. For the predictions of the OPTIMISTIC 10-month and Canadian cohort values using the baseline model, out-of-sample R-squared values are also reported, which were calculated based on:

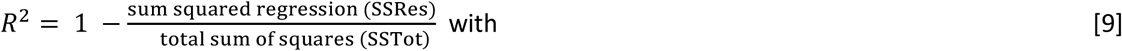

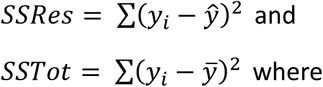

𝑦_𝑖_ = 𝑜𝑏𝑠𝑒𝑟𝑣𝑒𝑑 𝑡𝑒𝑠𝑡𝑖𝑛𝑔 𝑑𝑎𝑡𝑎, 𝑦^ = 𝑝𝑟𝑒𝑑𝑖𝑐𝑡𝑒𝑑 𝑤𝑖𝑡ℎ 𝑏𝑎𝑠𝑒𝑙𝑖𝑛𝑒 𝑚𝑜𝑑𝑒𝑙, 𝑦̅ = 𝑚𝑒𝑎𝑛 𝑜𝑓 𝑜𝑏𝑠𝑒𝑟𝑣𝑒𝑑 𝑡𝑒𝑠𝑡𝑖𝑛𝑔 𝑑𝑎𝑡𝑎

For the most optimal VIP value (60%), the coefficients and p-values of the associated multivariate OPTIMISTIC baseline model are reported (n=209). To assess linear model assumptions, univariate multiple regression models were generated to individually predict CTG-repeat (n=210) and 6MWT (n=212) scores based on the identified protein group set using the OPTIMISTIC baseline data. Linear model assumptions (Global Stat, Skewness, Kurtosis, Link Function, Heteroscedasticity) of these two models were checked using the gvlma package^43^. If assumptions were not met, influential datapoints were identified through visual inspection of Cook’s distances.

## Results

### Patient cohorts characteristics and comparisons

This study focused on the identification of protein biomarkers in serum samples of DM1 patients. For this purpose, bottom-up mass-spectrometry (MS)-based proteomic profiles of more than 400 serum samples of the OPTIMISTIC study cohort were generated. The OPTIMISTIC samples were derived from two study timepoints, at the start of the trial (n = 235) and the primary trial endpoint after 10 months (n = 202). OPTIMISTIC was not only the largest clinical trial in DM1 to date but also characterised by rich phenotype profiling of all patients. In addition to measuring the CTG-repeat expansion in blood, which we use as a proxy for both disease severity and pathology in this study, 25 different clinical outcome measures were used. We grouped all outcome measures into 6 disease domains which, including the abbreviations used in this study and the corresponding full names, are summarized in table 1. We refer the reader for to earlier publications for more details on how the clinical measurements were obtained and specific scoring ranges, including the calculation of interference scores such as those for the Stroop Colour-Word Test ^8,9^.

To externally validate the study findings, serum-based MS-based proteomic profiles were also generated for 56 Canadian DM1 patients, 13 German DM1 patients and 10 German healthy controls in an independent laboratory (Table 2). For all 69 DM1 patients, information on the CTG-repeat was present, and for the 56 DM1 patients from the Canadian cohort, also information on the six-minute walk test (6MWT) and grip strength of both hands was available.

**Table 2:**
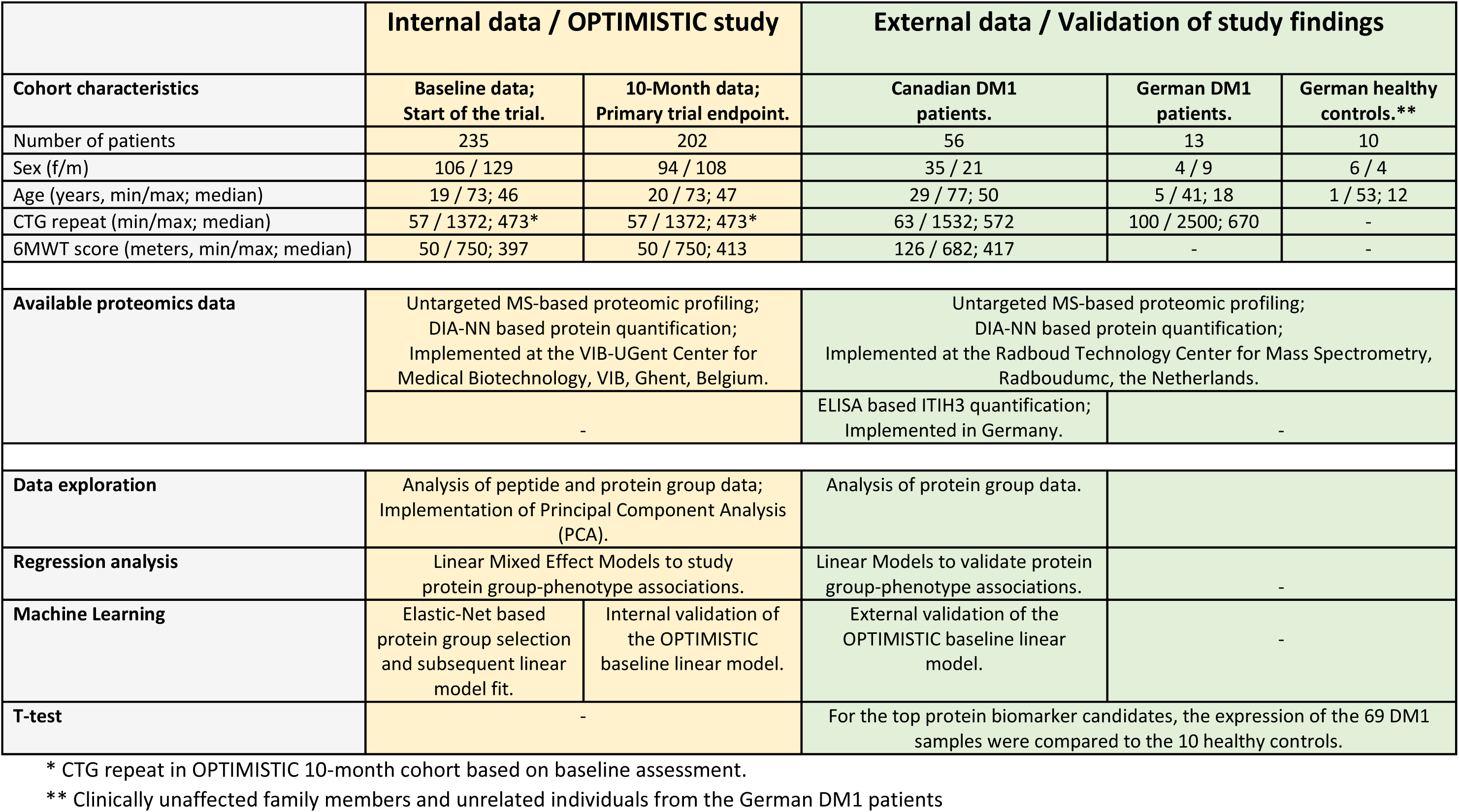
Overview of patient cohorts, proteomic datasets and implemented analyses.

|  | Internal data / OPTIMISTIC study |  | External data / Validation of study findings |  |  |
| --- | --- | --- | --- | --- | --- |
| Cohort characteristics | Baseline data;<br>Start of the trial. | 10-Month data;<br>Primary trial endpoint. | Canadian DM1<br>patients. | German DM1<br>patients. | German healthy<br>controls.** |
| Number of patients | 235 | 202 | 56 | 13 | 10 |
| Sex (f/m) | 106 / 129 | 94 / 108 | 35 / 21 | 4 / 9 | 6 / 4 |
| Age (years, min/max; median) | 19 / 73; 46 | 20 / 73; 47 | 29 / 77; 50 | 5 / 41; 18 | 1 / 53; 12 |
| CTG repeat (min/max; median) | 57 / 1372; 473* | 57 / 1372; 473* | 63 / 1532; 572 | 100 / 2500; 670 | - |
| 6MWT score (meters, min/max; median) | 50 / 750; 397 | 50 / 750; 413 | 126 / 682; 417 | - | - |
| Available proteomics data | Untargeted MS-based proteomic profiling;<br>DIA-NN based protein quantification;<br>Implemented at the VIB-UGent Center for<br>Medical Biotechnology, VIB, Ghent, Belgium. |  | Untargeted MS-based proteomic profiling;<br>DIA-NN based protein quantification;<br>Implemented at the Radboud Technology Center for Mass Spectrometry,<br>Radboudumc, the Netherlands. |  |  |
|  | - |  | ELISA based ITIH3 quantification;<br>Implemented in Germany. | - |  |
| Data exploration | Analysis of peptide and protein group data;<br>Implementation of Principal Component Analysis<br>(PCA). |  | Analysis of protein group data. |  |  |
| Regression analysis | Linear Mixed Effect Models to study<br>protein group-phenotype associations. |  | Linear Models to validate protein<br>group-phenotype associations. | - |  |
| Machine Learning | Elastic-Net based<br>protein group selection<br>and subsequent linear<br>model fit. | Internal validation of<br>the OPTIMISTIC<br>baseline linear model. | External validation of the<br>OPTIMISTIC baseline linear<br>model. | - |  |
| T-test | - |  | For the top protein biomarker candidates, the expression of the 69 DM1<br>samples were compared to the 10 healthy controls. |  |  |

\* CTG repeat in OPTIMISTIC 10-month cohort based on baseline assessment.
\*\* Clinically unaffected family members and unrelated individuals from the German DM1 patients

At both time points, the sex distribution was roughly balanced, and the age distributions were comparable. Based on the minimum, maximum and median values, the CTG-repeat length was also comparable between the two cohorts (Table 2). However, the distributions of the OPTIMISTIC CTG-repeat scores resembled a slightly-right skewed normal distribution. In contrast, the distribution of the Canadian cohort-based CTG-repeat scores was more uniform with a right skew (SFigure 1, panels A, B, C). Similar to the CTG-repeat, the minimum, maximum and median values of the 6MWT scores were also roughly comparable between the OPTIMISTIC and Canadian cohorts (Table 2). The 6MWT scores of the OPTIMISTIC cohort resemble a slightly right-skewed normal distribution, whereas the 6MWT scores of the Canadian cohort showed a more uniform distribution (SFigure 2, panels A, B, C).

**Figure 1:**
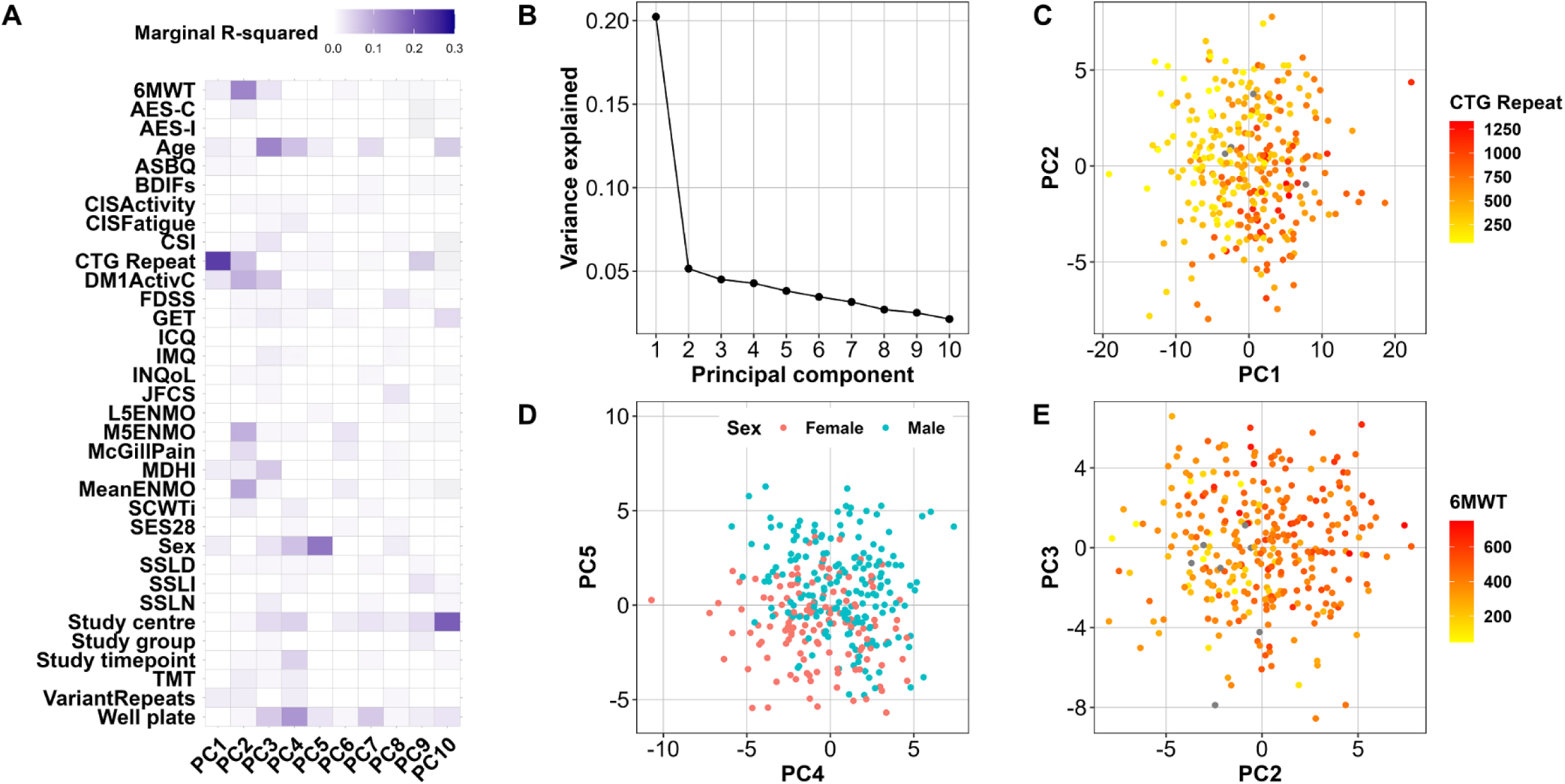
Drivers of protein variance in OPTIMISTIC serum samples. Results from PCA on a filtered subset of the OPTIMISTIC protein samples with complete observations. For the first ten principal components (PCs), a mixed-effect model was fitted with various phenotype measures as predictors and PatientID as a random effect (model 1). (A) Heatmap of the marginal R-squared values for each of the phenotype measures (rows) and the first 10 PCs (columns). (B) Scree plot showing the proportion of variance explained by the first 10 PCs. (C) Plot of sample scores on PC1 and PC2 coloured by CTG repeat length. (D) Plot of sample scores on PC4 and PC5 coloured by biological sex. (E) Plot of sample scores on PC2 and PC3 coloured by distance on the 6MWT. Grey dots reflect missing phenotype information. Abbreviations: See *Table 1* for outcome measures; GET Graded Exercise Therapy.

**Figure 2:**
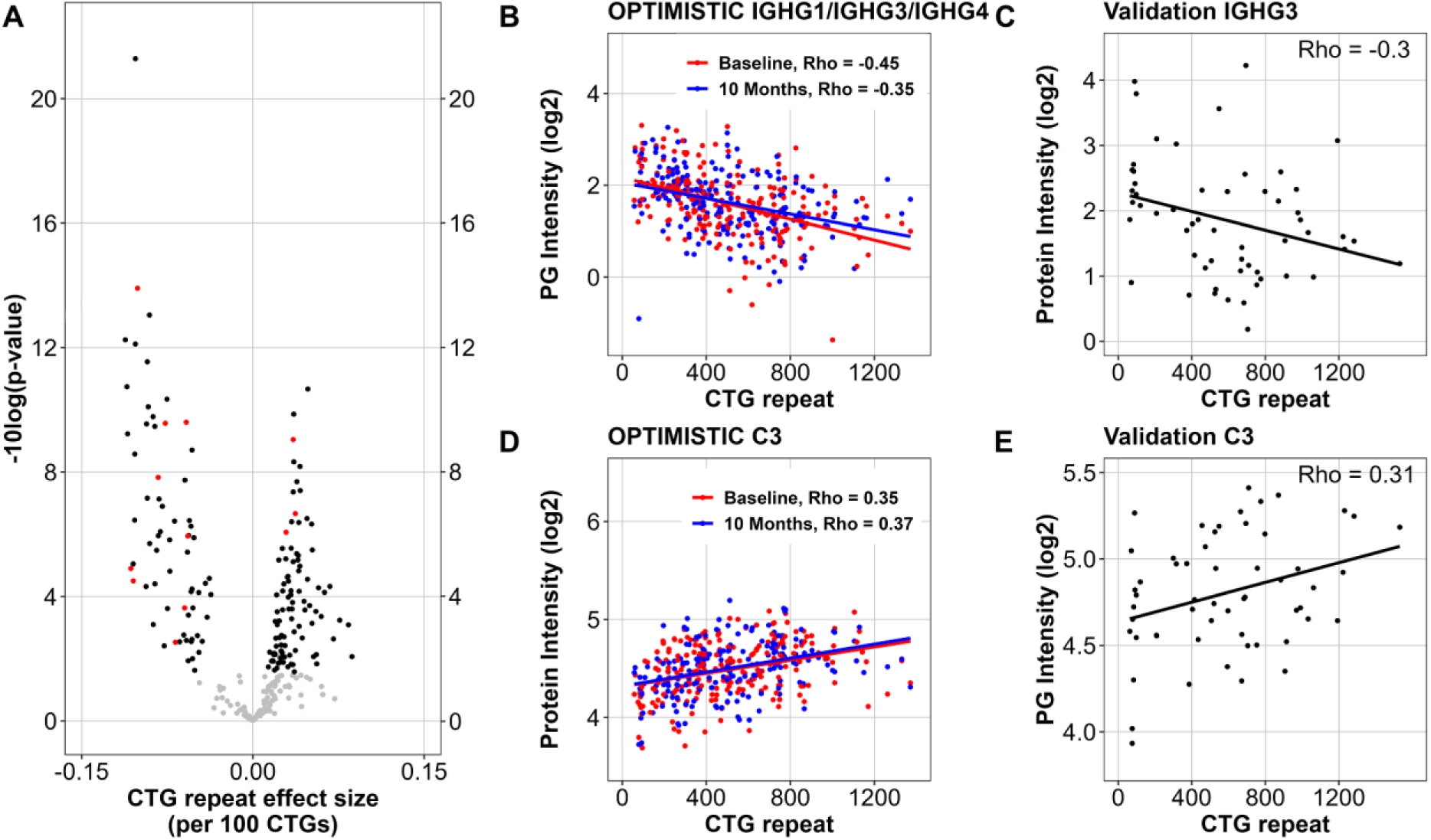
Protein group associations with CTG-repeat length. For each identified protein group (PG) in the OPTIMISTIC serum samples, a mixed effect model (model 2) was fitted with the CTG-repeat as predictor, the covariates sex and study timepoint as fixed and PatientID and well plate as random effects. (A) Volcano plot showing the significance (-log10 of nominal p-values, y-axis) and the effect size of the CTG-repeat (per 100 CTG repeats, x-axis) for the protein groups. Protein groups for which the CTG-repeat was significant (FDR < 0.05) are visualized in black, proteins that were also significant (p < 0.05) in the Canadian validation dataset are visualized in red. (B-C) Scatter plots of the abundance of the protein group IGHG1/IGHG3/IGHG4 and the protein IGHG3 (y-axis) plotted against the CTG-repeat in the OPTIMISTIC and external Canadian validation cohort, respectively. (D-E) Scatter plots of the abundance of the complement protein C3 (y-axis) plotted against the CTG-repeat in the OPTIMISTIC and external Canadian validation cohort, respectively. In Panels B and D, red dots represent samples taken at baseline, and blue dots represent samples taken 10 months after the start of the trial. Rho indicates Pearson’s correlation coefficient.

### Quality control

Since not all peptides could be uniquely matched with a protein, protein groups are reported instead. This group includes all possible protein matches that are equally likely to be present in the samples. After all filtering steps, a total of 259 protein groups were identified in both the OPTIMISTIC and Canadian cohorts. For the 189 paired OPTIMISTIC samples (from the same patient, at two timepoints), the Pearson correlation was calculated for each protein group between the baseline and the matched 10-month expression. This was compared to the average protein group correlation between the baseline and all other 10-month samples. The average correlation of samples from the same patient was 0.97, whereas the average correlation across all unrelated sample combinations was 0.94 (SFigure 3A). For 143 out of the 189 patients, the correlation of the baseline sample was higher with its matched 10-month sample than with any other 10-month sample in the dataset.

**Figure 3:**
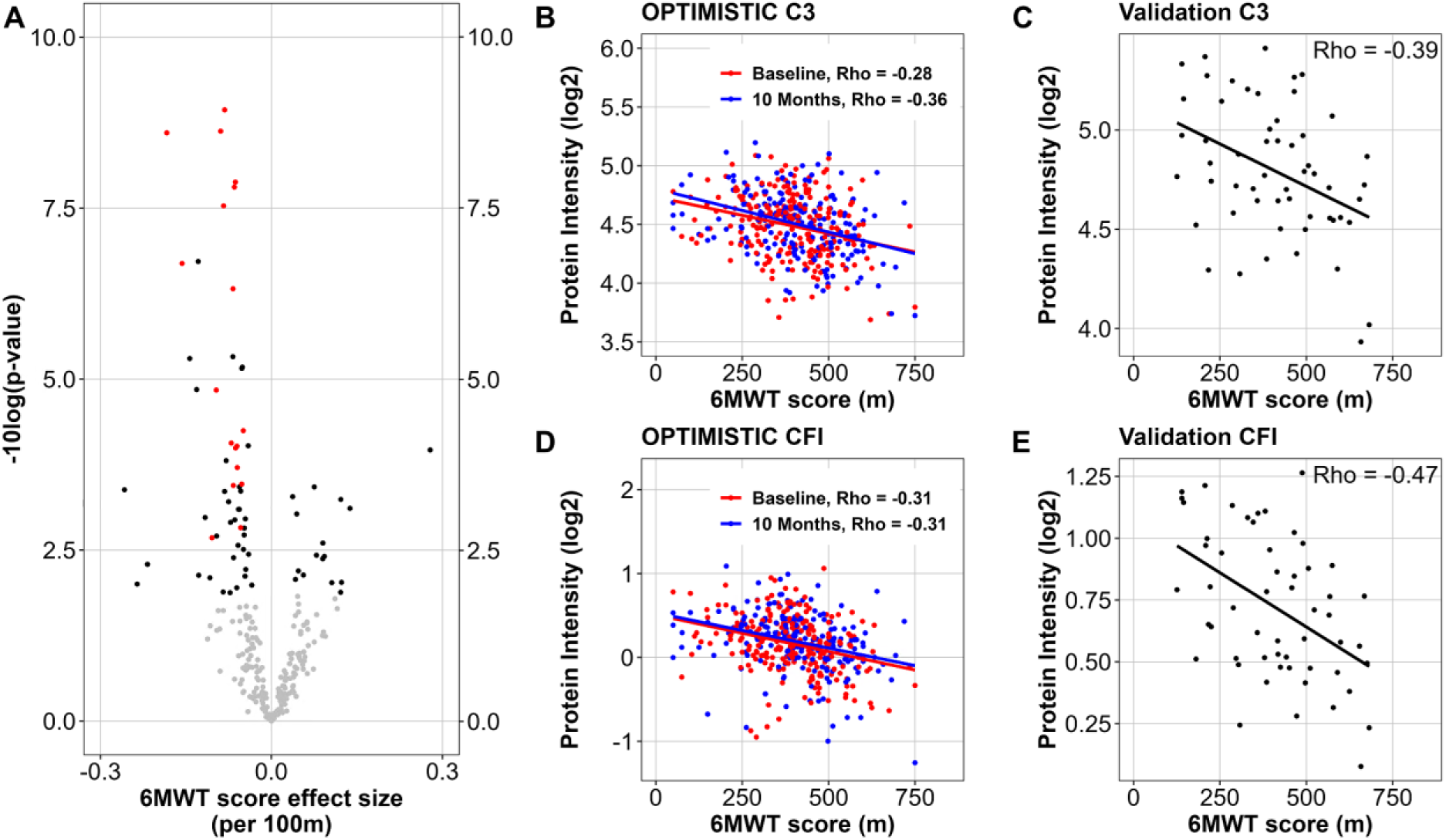
Protein group associations with the 6MWT score. *For each identified protein group in the OPTIMISTIC serum samples, a mixed effect model (model 3) was fitted with the 6MWT as predictor, the covariates sex and study timepoint as fixed and PatientID and well plate as random effects. (A) Volcano plot showing the significance (-log10 of nominal p-values (y-axis) and the effect size of the 6MWT for the protein groups (x-axis). Protein groups for which the 6MWT was significant (FDR < 0.05) are visualized in black, proteins that were also significant (p < 0.05) in the Canadian validation dataset are visualized in red. (B-E) Scatter plots of the abundance of Complement C3 (Panels B and C) and Complement factor I (CFI, Panels D and E) (y-axis) against the 6MWT score in metres (x-axis) in the OPTIMISTIC cohort (Panels B and D) and the external Canadian validation cohort (Panels C and E). In Panels B and D, red dots represent samples taken at baseline, and blue dots represent samples taken 10 months after the start of the trial. Rho indicates Pearson’s correlation coefficient*.

From this, we conclude that there were no systematic sample mix-ups. However, we cannot rule out the possibility of an occasional sample mix-up in our study. Mean-variance protein intensity plots of the OPTIMISTIC samples and the external validation samples demonstrated adequate data normalization and the expected pattern of slightly higher variation in low-abundant proteins (SFigure 3, panels C, D).

### The CTG-repeat length is the strongest driver of variance in protein abundance

Principal component analysis (PCA) was applied to identify which clinical variables are important drivers of differences in serum peptide and protein abundance within the OPTIMISTIC cohort. For the protein data, the first principal component (PC1) was most strongly associated with the CTG-repeat length, indicating that differences in CTG-repeat sizes can explain up to 20% of differences in the blood proteome in DM1 patients (Figure 1, panels A, B, C). Interestingly, PC2 was most strongly linked to the 6MWT (Figure 1, panels A, E). The association of PC5 with biological sex was an expected finding, as the blood proteome is known to differ between men and women (Figure 1, panels A, D)^44^. The PCA implemented on the peptide-level data revealed a surprising association between PC1 and the plate measured in the mass spectrometer (SFigure 4, Panel A, E). Further analysis suggests that this is likely due to an uneven distribution of study timepoints across the well plates, with baseline samples primarily located on plates 1–3 and 10-month samples predominantly on plates 4 and 5 (SFigure 3, panel B). Consequently, to correct for this imbalance, the plate effect was included as a random effect in the statistical analyses of the OPTIMISTIC data. Phenotype PC associations were generally weaker for the peptide data; however, the CTG-repeat was also a driver of variance here (associated with PC4 and PC5, SFigure 4, panels A, B, C). The association of biological sex with a PC was weaker for the peptide data as well, although a clear separation along PC8 and PC9 was found (SFigure 4, panel D). Given better interpretability and robustness, we continued with the analysis of the protein group-level data.

**Figure 4:**
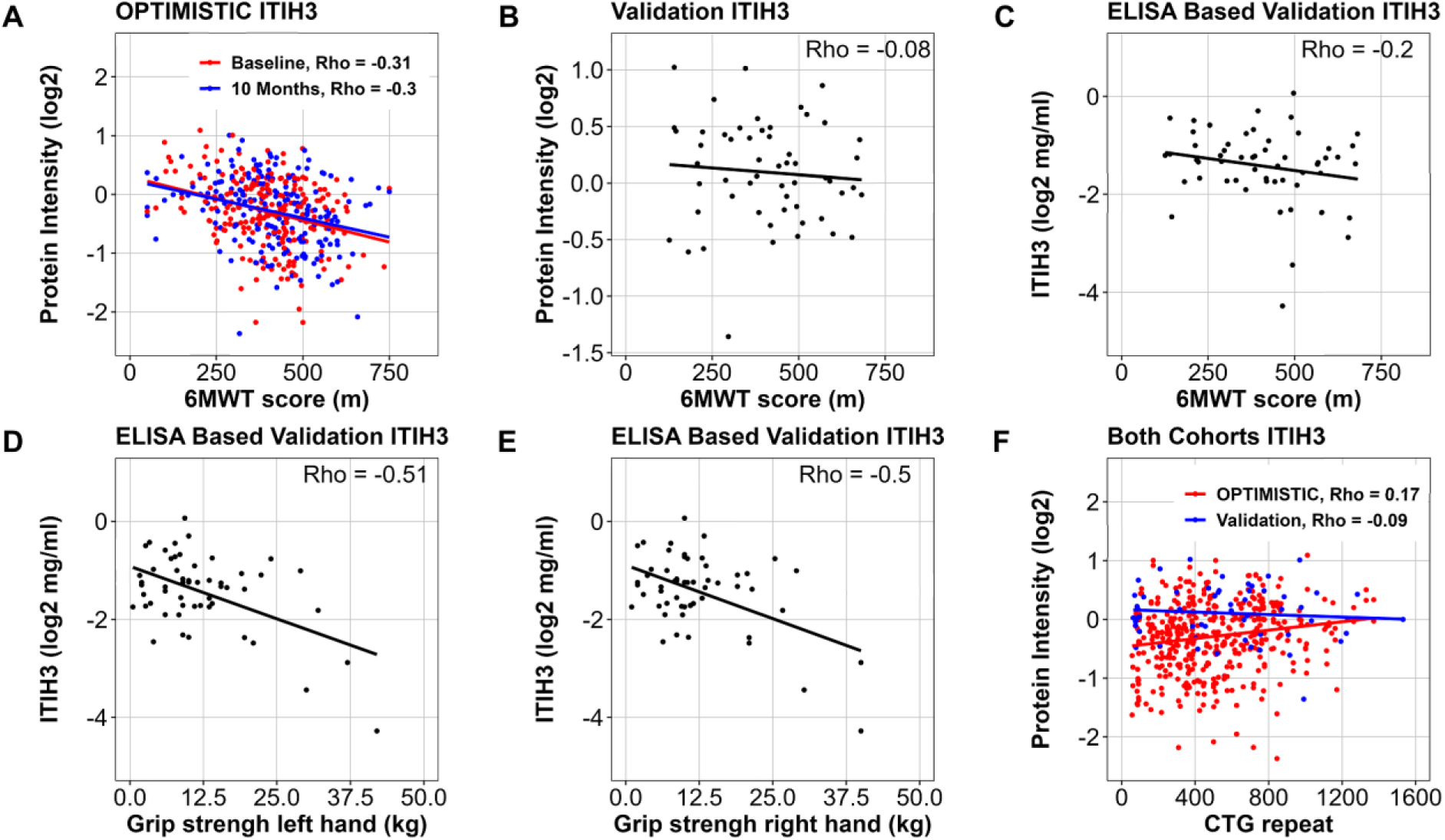
ITIH3 associations with markers of physical performance and CTG-repeat length. Association of the MS-quantified relative abundance levels of ITIH3 abundance with the 6MWT scores for the OPTIMISTIC samples (Panel A, colour coded by study visit) and external Canadian validation cohort (Panel B). Association of the ELISA-based quantification of the absolute ITIH3 abundance (mg/ml, log-scale) with the 6MWT scores (Panel C) and grip strength measures (left (Panel D) and right (Panel E) hand) in the external Canadian validation cohort samples. Association of the MS-quantified ITIH3 relative abundance levels with the CTG-repeat length in the OPTIMISTIC (red) and external Canadian validation (blue) cohorts (Panel F). Rho indicates Pearson’s correlation coefficient.

### Serum protein associations with CTG-repeat length confirm hypogammaglobulinemia in DM1

Out of the 259 protein groups discovered in the OPTIMISTIC serum samples, 161 were significantly associated with CTG-repeat length (FDR < 0.05; Figure 2, panel A). This finding is in line with the results from the PCA. The validity of this result is further supported by the finding that no significant associations remained after randomization of the CTG-repeat length values. Twelve of the identified candidate protein biomarkers were matched with protein groups that were also significantly associated with CTG-repeat length in the external Canadian validation cohort (p < 0.05; Figure 2, panel A; Table 3). Among these validated findings, the absolute strongest correlations with the CTG repeat were observed for immunoglobulin heavy constant gamma chains IGHG1, IGHG3 and IGHG4 (Table 3, Figures 2B and 2C). All of the significant and validated immunoglobulin-related protein groups were negatively associated with the CTG-repeat length, confirming hypogammaglobulinemia in DM1. Furthermore, based on the complete external validation dataset, the four immunoglobulin biomarkers with the strongest negative Pearson Rho value show a consistent trend of lower expression in DM1 patients compared to healthy controls (SFigure 5, panels A-D). In addition to the negative association between the CTG-repeat length and extent of the hypogammaglobulinemia, it is interesting to note that the CTG-repeat length was also positively associated with several protein groups of the complement system (C3, CFI, CFH, Table 3). Given the milder clinical phenotype in DM1 patients with an interruption of the CTG-repeat, we expected milder protein group-CTG repeat associations in these patients. However, similar associations with both immunoglobulins and complement factors were found for patients with a CTG repeat interruption (SFigure 6, panels A-F).

**Table 3:** Top significant and validated protein associations with CTG-repeat and 6MWT score.

| Association | OPTIMISTIC<br>Protein group ID<br>[Gene names] | Protein names | OPTIMISTIC<br>adjusted<br>p-value* | OPTIMISTIC<br>Pearson Rho | Canadian DM1 cohort<br>Matched Protein group ID<br>[Gene Names] | Canadian<br>DM1 cohort<br>p-value | Canadian<br>DM1 cohort<br>Pearson Rho |
| --- | --- | --- | --- | --- | --- | --- | --- |
| **CTG Repeat** | IGHG1;IGHG3;IGHG4 | Immunoglobulin heavy constant gamma 1; 3; 4 | < 0.0001 | -0.4078 | IGHG3 | 0.0343 | -0.2998 |
|  | C3 | Complement C3 | < 0.0001 | 0.3576 | C3 | 0.021 | 0.3147 |
|  | IGLL5 | Immunoglobulin lambda-like polypeptide 5 | < 0.0001 | -0.346 | IGLL5;IGLC1;IGL1;IGLC2;IGLC3 | 0.0111 | -0.3312 |
|  | IGLC7;IGLL5 | Immunoglobulin lambda constant 7 | < 0.0001 | -0.3371 | IGLC7;IGLL5;IGLC6;IGLC1;IGL1;IGLC2;IGLC3 | 0.0049 | -0.3664 |
|  | IGLV3-9;IGLV3-21 | Immunoglobulin lambda variable 3-9; | < 0.0001 | -0.3144 | IGLV3-9; IGLV3-21 | 0.0366 | -0.2959 |
|  | CFI | Complement factor I | < 0.0001 | 0.2955 | CFI | 0.0336 | 0.3027 |
|  | CFH | Complement factor H | < 0.0001 | 0.2899 | CFH | 0.0386 | 0.2583 |
|  | IGLV8-61 | Immunoglobulin lambda variable 8-61 | 0.0001 | -0.2883 | IGLV8-61 | 0.0223 | -0.3348 |
|  | IGHV3-74;IGHV3-7 | Immunoglobulin heavy variable 3-74; 3-7 | < 0.0001 | -0.2584 | IGHV3-21; IGHV3-74;IGHV3-66;IGHV7-4-1;<br>IGHV3-11;IGHV3-48;IGHV3-53;IGHV3-33;<br>IGHV3-7;IGA2;IGHV3-30-3 | 0.0477 | -0.2542 |
|  | IGHV1-69 | Immunoglobulin heavy variable 1-69 | 0.0001 | -0.2413 | IGHV1-69D;IGHV1-3;IGHV1-69;<br>IGHV1-46;IGHV1-8 | 0.0223 | -0.4992 |
|  | IGLC2;IGLC3 | Immunoglobulin lambda constant 2; 3 | 0.0007 | -0.1834 | IGLC2;IGLC3 | 0.0199 | -0.2951 |
|  | IGKV2-28;IGKV2D-28 | Immunoglobulin kappa variable 2-28; 2D-28 | 0.0061 | -0.1477 | IGKV2-28;IGKV2D-29;IGKV2D-30;<br>IGKV2-40;IGKV2D-26;IGKV2-29;<br>IGKV2D-40; IGKV2D-28;IGKV2-30 | 0.0277 | -0.2586 |
| **6MWT** | C3 | Complement C3 | < 0.0001 | -0.3148 | C3 | 0.0027 | -0.3946 |
|  | CFI | Complement factor I | < 0.0001 | -0.3071 | CFI | 0.0006 | -0.4704 |
|  | C5 | Complement C5 | < 0.0001 | -0.3065 | C5 | 0.007 | -0.3701 |
|  | LGALS3BP | Galectin-3-binding protein | < 0.0001 | -0.3009 | LGALS3BP | 0.0181 | -0.3924 |
|  | CFH | Complement factor H | < 0.0001 | -0.299 | CFH | 0.0004 | -0.4619 |
|  | HP;HPR | Haptoglobin; Haptoglobin-related protein | < 0.0001 | -0.288 | HP;HPR | 0.0215 | -0.3452 |
|  | C4BPA | C4b-binding protein alpha chain | < 0.0001 | -0.2877 | C4BPA | 0.0037 | -0.3975 |
|  | C9 | Complement component C9 | 0.0002 | -0.2522 | C9 | 0.0018 | -0.4273 |
|  | AMBP | Protein AMBP | < 0.0001 | -0.2287 | AMBP | 0.0079 | -0.3136 |
|  | C6 | Complement component C6 | 0.0022 | -0.2282 | Complement component C6 | 0.0037 | -0.3642 |
|  | LCAT | Phosphatidylcholine-sterol acetyltransferase | 0.0013 | -0.2165 | LCAT | 0.0101 | -0.3582 |
|  | SERPINA3 | Alpha-1-antichymotrypsin | 0.0036 | -0.2138 | SERPINA3 | 0.007 | -0.3815 |
|  | C2 | Complement C2 | 0.0091 | -0.1994 | C2 | 0.0234 | -0.3196 |
|  | C1R | Complement C1r subcomponent | 0.0009 | -0.1894 | C1R;C1RL | 0.0034 | -0.424 |
|  | C4BPB | C4b-binding protein beta chain | 0.0013 | -0.1856 | C4BPB | 0.0025 | -0.3729 |
|  | F9 | Coagulation factor IX | 0.0036 | -0.1765 | F9 | 0.0014 | -0.39 |
|  | LBP | Lipopolysaccharide-binding protein | 0.0118 | -0.1705 | LBP | 0.0321 | -0.3205 |
|  | APCS | Serum amyloid P-component | 0.0013 | -0.1418 | APCS | < 0.0001 | -0.4662 |

\* FDR based on Benjamini-Hochberg's multiple testing correction procedure

### Increased abundance of complement factors in DM1 patients with reduced functional capacity

When studying the associations of protein groups with individual clinical outcome measures, a surprising pattern emerged. In line with the PCA findings, the majority of FDR corrected significant associations were linked to measures of functional capacity (6MWT, DM1ActivC, Actometry based activity; Table 4). Yet virtually no other associations were found with disease domains such as fatigue, social functioning, cognitive function, disease impact or apathy. The most significant associations were found for the 6MWT score (n=70; Table 4; Figure 3, panel A), encompassing most of the hits found with DM1ActivC (45 out of 61), MeanENMO (29 out of 36) and M5ENMO (28 out of 32). The finding further supports the validity of the associations with the 6MWT scores, as only one significant association was found after randomizing the 6MWT scores.

**Table 4:** Numbers of significant protein group associations with clinical outcome measures.

| **Functional Capacity** | **p-value**<br>**< 0.05** | **FDR**<br>**< 0.05** | **Cognitive function,**<br>**Depression, Illness Coping** | **p-value**<br>**< 0.05** | **FDR**<br>**< 0.05** |
| --- | --- | --- | --- | --- | --- |
| 6MWT | 100 | 70 | SES28 | 51 | 0 |
| DM1ActivC | 106 | 61 | IMQ | 26 | 0 |
| MeanENMO | 73 | 36 | SCWTi | 25 | 0 |
| M5ENMO | 63 | 32 | ICQ | 21 | 0 |
| CISActivity | 40 | 0 | TMT | 15 | 0 |
| L5ENMO | 8 | 0 | BDIFs | 9 | 0 |
| **Fatigue** |  |  | **Disease Impact, Severity and Pain** |  |  |
| FDSS | 22 | 2 | MDHI | 70 | 23 |
| JFCS | 21 | 0 | INQoL | 50 | 1 |
| CISFatigue | 17 | 0 | McGillPain | 22 | 1 |
| **Social Functioning, Behaviour**<br>**and Impact on Caregivers** |  |  | **Apathy** |  |  |
| SSLN | 34 | 2 | AES-C | 31 | 0 |
| CSI | 23 | 0 | AES-I | 8 | 0 |
| SSLI | 23 | 0 |  |  |  |
| ASBQ | 18 | 0 |  |  |  |
| SSLD | 8 | 0 |  |  |  |

Out of the 70 protein groups associated with the 6MWT, 18 were matched with protein groups that were also significant in the external Canadian validation cohort (Table 3, Figure 3, panel A).

Interestingly, the majority of these 18 validated hits were also significantly associated with DM1ActivC (n=17, excluding LBP), MeanENMO (n=17, excluding LBP) and M5ENMO (n=15, excluding LBP, SERPINA3 and HPT;HPR) in the OPTIMISTIC cohort. Among these, the two protein groups that show the strongest absolute correlation with the 6MWT are complement C3 and complement factor I, which exhibit an increase in abundance in patients with reduced functional capacity (Table 3; Figure 3, panels B-E). This increase is also observed for all other validated complement components or factors (Table 3). The abundance of the top biomarker candidates (complement C3 and C5, complement factor I) also showed a trend of higher expression in DM1 patients compared to healthy controls (SFigure 5, panels E-G).

In the OPTIMISTIC cohort, one of the significant findings with the 6MWT was ITIH3 (inter-alpha-trypsin inhibitor heavy chain 3), which has previously been linked to complement modulation and disease severity in Myasthenia Gravis (FDR < 0.001; Figure 4, panel A)^45^. While this negative association with the 6MWT could neither be validated in the MS-nor ELISA-based protein quantification of the external Canadian cohort (resp. p=0.4, p= 0.21, Figure 4, panels B, C), external significant negative associations were found for the ELISA-based protein quantification and grip strength of both the left and right hand (both p < 0.001; Figure 4, panels D, E). Interestingly, ITIH3 showed a significant weak positive association with the CTG-repeat size in the OPTIMISTIC cohort (FDR = 0.014), which was, however, not validated in the MS-based protein quantification of the Canadian cohort (p = 0.58) (Figure 4, panel F). Based on the complete external validation cohort, no difference in ITIH3 expression was found between DM1 patients and healthy controls (SFigure 5, panel H).

### No effect of Cognitive Behavioural Therapy or Graded Exercise on the serum blood proteome

Given the significant clinical effects of Cognitive Behavioural Therapy (CBT) on the ability to perform daily tasks and to participate in social activities in the OPTIMISTIC clinical trial, we investigated whether these benefits also translated to significant changes in the serum proteome of DM1 patients. However, only eight significant changes between the CBT and standard-of-care group were identified (Vitamin D-binding protein, Complement C5 and C9, Ceruloplasmin, Immunoglobulin heavy constant gamma 3 and mu, Inter-alpha-trypsin inhibitor heavy chain H4, Carboxypeptidase B2; SFigure 7, panels A-I). Upon closer inspection, none showed a convincing link with the CBT intervention, and most were associated with changes in the standard-of-care group over the 10-month study period. Since a subgroup of patients in the intervention group also completed a graded exercise program (GET), we also investigated whether proteome changes could be linked to this intervention. Yet only two proteins were significantly linked to GET (Cadherin-5, Fibronectin; SFigure 8, panel A). Upon closer inspection, these proteins did not show convincing differences between the study groups either (SFigure 8, panels B-C). Since samples were not evenly distributed across well plates for study timepoint (SFigure 3, panel B), the random effect associated with the ‘well plate’ variable may have confounded this finding. However, applying the same statistical frameworks without the random plate effect did not yield additional findings for either CBT or GET.

### Combined set of proteins associated with both CTG-repeat length and 6MWT score

We subsequently tested whether a set of protein groups would show stronger associations than individual biomarkers. Since the ideal set of biomarkers is strongly associated with both the disease pathology (disease relevant) and the clinical phenotype (clinically relevant), a machine learning based algorithm (bootstrap enhanced Elastic-Net regression) was implemented to find a minimum subset of proteins that together optimally explain the variance of both the CTG-repeat length (disease pathology) and 6MWT scores (clinical phenotype). As a starting point, we considered 47 protein groups that were significantly associated with both the CTG-repeat length (model 2) and the 6MWT score (model 3). We subsequently selected the 42 protein groups that were also unambiguously identified in the external Canadian validation dataset. In an effort to harmonize the distributional differences for the CTG-repeat length and 6MWT scores between the OPTIMISTIC and external Canadian validation dataset, these two dependent variables were box-cox transformed to more closely resemble a normal distribution, and subsequently standardized (SFigure1, panels D-F; SFigure 2, panels D-F). For each of the 42 protein groups, a Variable Inclusion Probability (VIP) score was generated based on how frequently the protein group was selected as a predictor for both CTG-repeat and 6MWT score across 50,000 statistical model fits (10 imputed datasets * 5,000 bootstrap distributions). These VIP scores were purely based on the OPTIMISTIC baseline dataset, and their predictive value was subsequently evaluated on the OPTIMISTIC 10M (internal validation, test set) and the external Canadian validation cohort (external validation set) samples (Table 5).

**Table 5:** Statistical performance estimates of multivariate predictions.

| Outcome | VIP (%) | Number of Predictors | Train RSQ | Train a. RSQ | Train RMSE | Test RSQ | Test RMSE | Val RSQ | Val RMSE |
| --- | --- | --- | --- | --- | --- | --- | --- | --- | --- |
| CTG | 90 | 5 | 0.2382 | 0.2213 | 0.872 | 0.1887 | 0.9003 | 0.0972 | 0.9274 |
|  | 80 | 10 | 0.318 | 0.2858 | 0.8195 | 0.2544 | 0.8632 | -0.0287 | 0.9863 |
|  | 70 | 12 | 0.348 | 0.3083 | 0.7932 | 0.2734 | 0.8416 | 0.1566 | 0.8779 |
|  | 60 | 13 | 0.3588 | 0.3162 | 0.79 | 0.2903 | 0.8318 | 0.1734 | 0.8692 |
|  | 50 | 18 | 0.3718 | 0.3126 | 0.782 | 0.2645 | 0.8513 | 0.2072 | 0.8512 |
| 6MWT | 90 | 5 | 0.205 | 0.1873 | 0.8932 | 0.2179 | 0.883 | 0.1476 | 0.981 |
|  | 80 | 10 | 0.3225 | 0.2905 | 0.8237 | 0.2861 | 0.8499 | 0.1719 | 0.9469 |
|  | 70 | 12 | 0.3201 | 0.2786 | 0.8178 | 0.2588 | 0.8362 | 0.2629 | 0.9165 |
|  | 60 | 13 | 0.3208 | 0.2757 | 0.8176 | 0.2561 | 0.8377 | 0.2565 | 0.9205 |
|  | 50 | 18 | 0.3392 | 0.2769 | 0.8067 | 0.2759 | 0.8305 | 0.3264 | 0.8761 |

\* VIP = Variable Inclusion Probability
\*\* Train = OPTIMISTIC baseline dataset; Test = OPTIMISTIC 10M dataset; Val = Validation dataset
\*\*\* RSQ = R-squared; a. RSQ = Adjusted R-squared; RMSE = Root Mean Square Error

We observed that up to 32% (Adjusted R-squared in the OPTIMISTIC training set) of the variance in CTG-repeat length can be explained with a set of only 13 protein groups (Table 5). When evaluating the performance of these protein groups in the internal and external validation data, we see a (slightly) lower amount of variance explained (respective R-squared values of 29 and 17%).

Interestingly, the same set of 13 protein groups can also explain up to 28% of the variance in the 6MWT scores of the OPTIMISTIC training set (Adjusted R-squared), while also showing similar performance for the internal and external validation data (both R-squared values are 26%). The 13 protein groups, together with the regression coefficient estimates for both the CTG-repeat and 6MWT scores of the OPTIMISTIC baseline model are summarized in Table 6. Univariate prediction models of the CTG-repeat and 6MWT with these 13 protein groups satisfied all assumptions of linear regression as assessed by the R-package gvlma after removal of one outlier sample identified through visual inspection of Cook’s distances^43^.

**Table 6:**
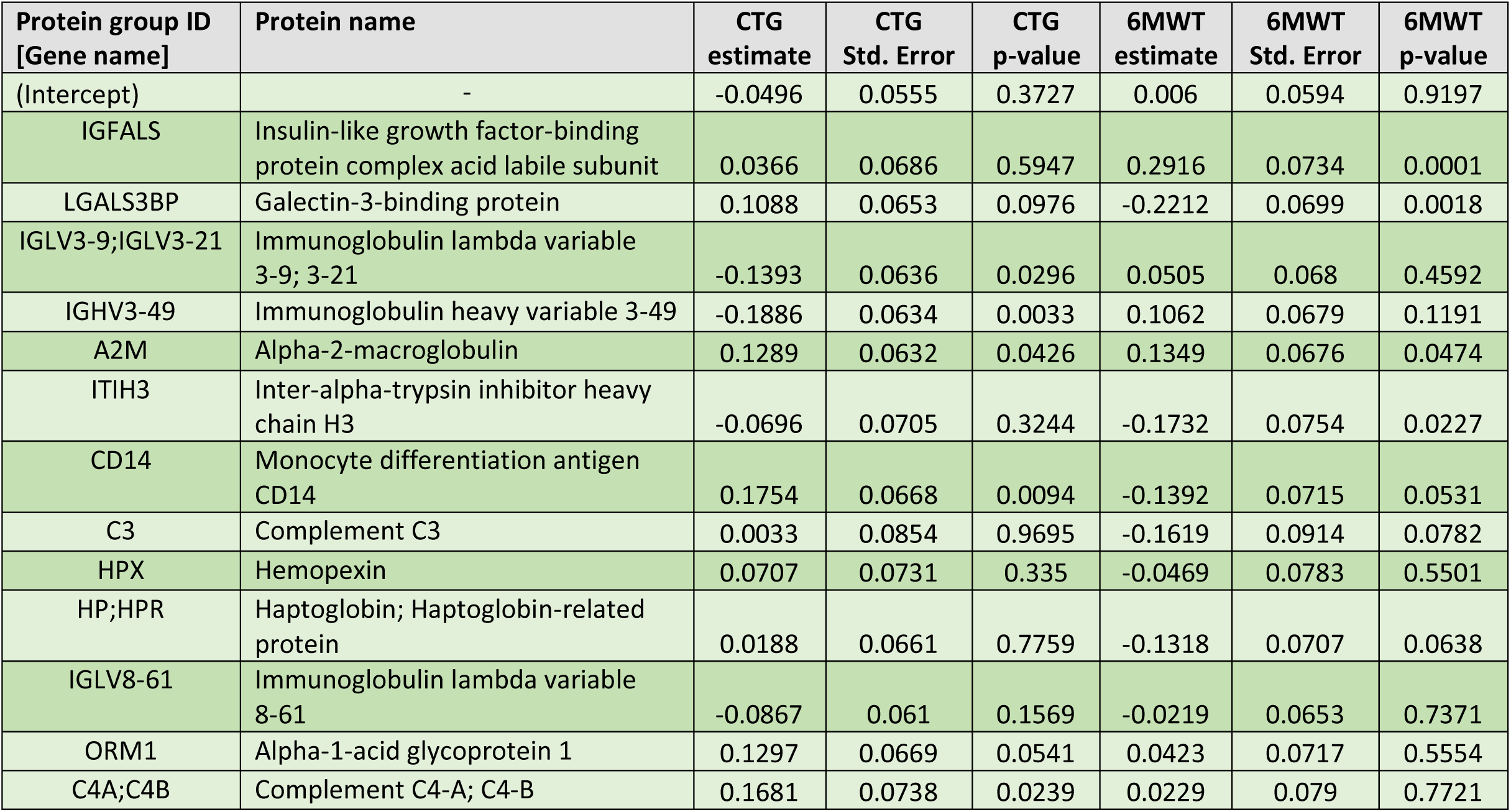
Coefficient estimates of multivariate OPTIMISTIC baseline CTG-repeat and 6MWT prediction.

| Protein group ID<br>[Gene name] | Protein name | CTG<br>estimate | CTG<br>Std. Error | CTG<br>p-value | 6MWT<br>estimate | 6MWT<br>Std. Error | 6MWT<br>p-value |
| --- | --- | --- | --- | --- | --- | --- | --- |
| (Intercept) | - | -0.0496 | 0.0555 | 0.3727 | 0.006 | 0.0594 | 0.9197 |
| IGFALS | Insulin-like growth factor-binding protein complex acid labile subunit | 0.0366 | 0.0686 | 0.5947 | 0.2916 | 0.0734 | 0.0001 |
| LGALS3BP | Galectin-3-binding protein | 0.1088 | 0.0653 | 0.0976 | -0.2212 | 0.0699 | 0.0018 |
| IGLV3-9;IGLV3-21 | Immunoglobulin lambda variable 3-9; 3-21 | -0.1393 | 0.0636 | 0.0296 | 0.0505 | 0.068 | 0.4592 |
| IGHV3-49 | Immunoglobulin heavy variable 3-49 | -0.1886 | 0.0634 | 0.0033 | 0.1062 | 0.0679 | 0.1191 |
| A2M | Alpha-2-macroglobulin | 0.1289 | 0.0632 | 0.0426 | 0.1349 | 0.0676 | 0.0474 |
| ITIH3 | Inter-alpha-trypsin inhibitor heavy chain H3 | -0.0696 | 0.0705 | 0.3244 | -0.1732 | 0.0754 | 0.0227 |
| CD14 | Monocyte differentiation antigen CD14 | 0.1754 | 0.0668 | 0.0094 | -0.1392 | 0.0715 | 0.0531 |
| C3 | Complement C3 | 0.0033 | 0.0854 | 0.9695 | -0.1619 | 0.0914 | 0.0782 |
| HPX | Hemopexin | 0.0707 | 0.0731 | 0.335 | -0.0469 | 0.0783 | 0.5501 |
| HP;HPR | Haptoglobin; Haptoglobin-related protein | 0.0188 | 0.0661 | 0.7759 | -0.1318 | 0.0707 | 0.0638 |
| IGLV8-61 | Immunoglobulin lambda variable 8-61 | -0.0867 | 0.061 | 0.1569 | -0.0219 | 0.0653 | 0.7371 |
| ORM1 | Alpha-1-acid glycoprotein 1 | 0.1297 | 0.0669 | 0.0541 | 0.0423 | 0.0717 | 0.5554 |
| C4A;C4B | Complement C4-A; C4-B | 0.1681 | 0.0738 | 0.0239 | 0.0229 | 0.079 | 0.7721 |

## Discussion

This study identified and externally validated a large set of candidate protein biomarkers in the serum of DM1 patients. Low-invasive biomarkers to monitor the response to treatments currently in development for DM1 are much needed, as they may demonstrate a faster and more homogeneous response compared to clinical outcome measures^46^.

The serum appeared to be a rich source of low-invasive disease-relevant biomarkers. This is highlighted by the finding that the strongest driver of variance in protein group expression (PC1) was linked to the CTG-repeat length, and that 161 out of the 259 protein groups were significantly associated with the CTG-repeat in the OPTIMISTIC cohort. Most importantly, twelve of these significant associations were confirmed in an independent set of 56 Canadian cohort samples measured in a different laboratory on a different MS instrument. This much smaller validated set of protein groups is most likely the result of the substantially smaller cohort size, leading to a much lower statistical power to detect and validate weaker associations. The strongest association was found with a protein group matched with IGHG1;IGHG3;IGHG4, confirming the known hypogammaglobulinemia in DM1. Although hypogammaglobulinemia in DM1 has been known for a long time, its underlying mechanism remains poorly understood. Serum comparisons of DM1 patients versus healthy controls have shown that the differences predominantly affect total IgG, IgG1, and IgG3, with no differences observed for other immunoglobulins (IgA, IgM, IgG2 and IgG4)^15,17^. Our results demonstrate that hypogammaglobulinemia is worse in more severely affected patients with longer CTG-repeats, a finding which has been reported before but was not consistently found in all studies^15,17,47^. Rather than a deficient production, hypogammaglobulinemia in DM1 appears to be caused by a more rapid breakdown of immunoglobulins, with one study also suggesting the possibility of extravascular redistribution due to increased capillary permeability^17,48,49^. While the exact mechanism of this disease-relevant finding remains unknown, the confirmed hypogammaglobulinemia strongly supports the validity of the untargeted MS-based approach used in this study for biomarker discovery.

A novel finding of our biomarker study is the elevation of protein groups associated with several components of the complement system in DM1 patients. Not only was this elevation positively correlated with the CTG-repeat length, but also with clinical scores reflecting reduced functional capacity. Moreover, the top immunoglobulins and complement components or factors both exhibited consistent patterns of respective down- and up-regulation in DM1 patients compared to healthy controls. While consistent, these findings must be interpreted with caution, given the small and heterogeneous cohort of the control group, particularly regarding age. The implemented MS workflow can only measure the abundance of complement proteins but cannot distinguish between unactivated (intact) and activated (cleaved) complement components. Complement activation markers arise from proteolytic cleavage, but the resulting peptides are indistinguishable from those of unactivated proteins, since the trypsin enzyme used in sample preparation cleaves at the same sites. Therefore, higher levels of complement proteins do not necessarily indicate increased complement activity but could also reflect increased hepatic production or reduced consumption of complement proteins. In addition to central complement components (C3, C5), several important regulators of the complement system, such as Complement Factor I and H, show a similar increase in abundance. In case of complement activation, this would suggest a controlled form of chronic inflammation. Complement activation is present in many chronic conditions such as type 2 diabetes, obesity and autoimmunity, all known comorbidities for DM1^50,51^.

We can only speculate on the exact causes of the potential complement activation in DM1. It may be due to a more general pro-inflammatory status in DM1 patients^52,53^. On the other hand, reduced serum levels of interleukin 6 (IL-6), a key regulator of the acute phase response and hepatic complement protein synthesis, have been previously shown to significantly correlate with muscle weakness and functional capacity limitations in DM1^54^. Further supporting the association with muscle pathology, the complement system is known to contribute to both fibrotic tissue remodelling and muscle fibre necrosis, and monitoring muscle fibrosis through serum periostin has recently been proposed as a novel stratification biomarker for DM1^55–57^. Another notable finding was the significant negative association between ITIH3 levels and 6MWT performance in the OPTIMISTIC cohort, which was, however, only partially and indirectly validated by significant negative ELISA-based correlations between ITIH3 and grip strength. ITIH3 has recently been identified as a potential biomarker for disease activity in Myasthenia Gravis (MG)^45^. Given the proposed role of ITIH3 in the early stages of complement activation, it has been hypothesized that elevated levels may be the result of an enhanced negative feedback loop in response to dysregulated complement activation. Considering the broad biological functions and disease associations of ITIHs, the authors further concluded that it is not a disease-specific marker, but rather has a potential use in already diagnosed patients to monitor disease activity. Our study in DM1 patients supports this conclusion. Furthermore, in addition to MG, dysregulation of the complement system has also been associated with other neuromuscular disorders such as Facioscapulohumeral muscular dystrophy (FSHD), Amyotrophic Lateral Sclerosis (ALS), and Duchenne Muscular Dystrophy (DMD)^58–60^. Even more so, modulators of the complement system are an active field of study and may provide valuable disease-modifying treatments within the broader neuromuscular field^59,60^.

Given the independent associations of the complement proteins with markers for functional capacity, one may also hypothesize that these changes are not exclusively disease associated. Also within the healthy populations, clear associations are described between higher fitness levels and decreased C3 levels in blood, as well as increased C1q levels with reduced muscle strength^61^. As a consequence, the recently demonstrated positive clinical effects of exercise in DM1 may also act as an independent disease modifier by attenuating complement activation^62–64^. Although some patients in the OPTIMISTIC intervention group also participated in a graded exercise therapy (GET) program, this unexpectedly had no significant effect on the serum proteome. This GET program was delivered to only a small number of patients in a highly individualized manner and did not yield clinical benefits beyond those achieved with cognitive behavioural therapy (CBT) alone^8^. We therefore hypothesize that a more standardized training program in a larger cohort is necessary to detect meaningful clinical improvement that is linked to complement modulation.

Despite the large number of protein groups being associated with markers of functional capacity, virtually no associations were found with other important DM1 disease-relevant domains such as apathy, cognition, fatigue, pain or social measures. While the blood-brain-barrier may prevent the detection of specific disease-associated proteins from the central nervous system (CNS) in serum, another well-studied CNS-derived biomarker, Neurofilament Light chain, is detectable in blood^65^. More sensitive and targeted detection methods quantifying absolute protein abundance may be necessary to identify molecular biomarkers associated with neurocognitive phenotypes. Considering the significant improvements of the DM1-Activ-C and the 6MWT scores in the OPTIMISTIC study, in combination with the abundant number of significant associations between protein groups and these outcome measures, it was surprising that no CBT-induced effects were observed in the blood proteome^8^. This was particularly surprising given that a very large number of genes were associated with the average intervention response in our previous transcriptomic study^14^. A confounder in the proteomic study was the uneven distribution of the OPTIMISTIC samples across the MS plates with regard to the study time point. Since this MS plate effect was regressed out, possible CBT-induced effects might have also been masked. Yet, peptide-level quantification seemed to suffer more from technical plate effects than protein-level quantification, and a supplemental analysis without regressing out the well plate effect did not reveal significant associations either. On the other hand, the general lack of overlap in identified biomarkers between our transcriptomic and proteomic study is likely the result of several biological and technical reasons. On a biological level, many serum proteins are either produced in the liver or secreted by various tissues and organs, whereas blood-based RNA-seq mostly reflects the leukocyte transcriptome. On a technical level, many of the mRNA biomarkers code for low-abundant proteins like cytokines, which are not detectable by our current MS-based methods.

Given that the significant associations with both the CTG-repeat and the 6MWT were individually relatively weak, we hypothesized that the clinical utility of these candidate biomarkers could be improved by finding a minimum combined subset of protein groups. Moreover, by combining multiple proteins, random variation in individual expression levels is more likely to average out, making the biomarker set a more robust indicator of disease state than individual proteins. The implemented bootstrap-enhanced Elastic-Net algorithm has robustly led to the identification of 13 protein groups that together can explain up to 32% and 28% of the variance of the CTG-repeat and 6MWT, respectively, while also performing comparatively well on the internal validation data. It is crucial to interpret the internal validation results with caution, as true independence is not achieved because many measurements originate from the same patients at different time points. Yet, even for the external validation, up to 17% and 26% of the CTG-repeat and 6MWT variance, respectively, were explained with our OPTIMISTIC baseline model. Given the inherent variability in protein expression, as well as for the CTG-repeat and 6MWT measurements, the amount of variance explained is in line with the expectations and supports the use of a combined set of proteins over individual proteins for therapeutic biomarker discovery. To establish their utility in clinical trials, further research is needed on longitudinal and potentially non-linear relationships between changes in the blood proteome and clinical outcomes.

## Conclusions

Our study extends the repertoire of lab-based biomarkers, including mRNA biomarkers and protein biomarkers, for potential use as surrogate endpoints in DM1 trials^14,65,66^. We have performed careful internal and external validation to confirm the robustness of the identified protein biomarkers. Going beyond individual protein associations, we demonstrated that a set of proteins is most likely to meet the statistical criteria required for surrogate clinical trial endpoints. Further longitudinal studies are needed to validate these findings, and methods that enable the absolute quantification of selected proteins are essential to advance their clinical utility.

## Data Availability

Researchers interested in accessing the phenotype data from the OPTIMISTIC study are invited to contact K. Mul (karlien.mul-at-radboudumc.nl) and complete a data access agreement. All requests will be reviewed by a panel comprising from each of the four participating clinical sites, with K. Mul serving as chair. The mass spectrometry proteomics data of the OPTIMISITIC samples have been deposited to the ProteomeXchange Consortium via the PRIDE partner repository with the dataset identifier PXD067476.
The phenotype data used and/or analysed from the Canadian cohort are available from the corresponding author upon reasonable request following the proper evaluation of the research protocol by the Ethics Review Board of the Centre intégré universitaire de santé et de services sociaux du Saguenay - Lac-St-Jean (Saguenay, Québec, Canada; cynthia.gagnon4 -at- usherbrooke.ca).
The phenotype data used from the German cohort, including the ELISA-based quantification data of ITIH3, are stored at the Department of Neurology of the University Hospital Duesseldorf (Heinrich Heine University) and are available upon request to roos -at andreas-roos.de. The mass spectrometry proteomics data of both the Canadian and German cohort have been deposited to the ProteomeXchange Consortium via the PRIDE partner repository with the dataset identifier PXD060035.

## Acknowledgements

We are grateful to the John Walton Muscular Dystrophy Research Centre Biobank, and in particular to Dan Cox, for the assistance in the storage and delivery of the patient samples used in this study. We would also like to thank dr. Dick Thijssen, Professor of Cardiovascular Physiology at the Radboudumc, for critically reviewing the manuscript.

OPTIMISTIC consortium: K. Okkersen, C. Jimenez-Moreno, S. Wenninger, F. Daidj, J. C. Glennon, S. Cumming, R. Littleford, D.G. Monckton, H. Lochmüller, M. Catt, C.G. Faber, A. Hapca, P.T. Donnan, G. Gorman, G. Bassez, B. Schoser, H. Knoop, S. Treweek, B.G.M. van Engelen.

ReCognitION consortium: D. van As, P.A. C. ‘t Hoen, D. G. Wansink, F. Impens, R. Gabriels, T. Claeys, A. Ravel-Chapuis, B.J. Jasmin, J.C. Glennon, N. Mahon, S. Nieuwenhuis, L. Martens, P. Novak, D. Furling, B.G.M. van Engelen, M. Catt, A. Baak, G. Gourdon, A. MacKenzie, C. Martinat, N. Neault, A. Roos, E. Duchesne, R. Salz, R. Thompson, S. Baghdoyan, A.M. Varghese, P. Blom, S. Spendiff, A. Manta.

## Author’s contributions

**D. van As:** Methodology, Validation, Software, Data Curation, Formal analysis, Writing – Original Draft, Writing – Review & Editing, Visualization. **T. Caeys, R. Salz, D. Van Haver, S. Dufour, A. van Deelen, J. Gloerich, R. Gabriels, P.J. Volders, A. van Gool, F. Impens,** and **L. Martens** Investigation, Resources, Data Curation, Writing – Review & Editing. **V. Dobelmann, A. Gangfuss, T. Ruck, G. Gourdon, E. Duchesne, C. Gagnon, A. Roos** Investigation, Resources, Writing – Review & Editing, Validation. **H. Lochmüller, B. Schoser, G. Bassez, B.G.M. van Engelen** Conceptualization, Writing – Review & Editing. **P.A.C. ‘t Hoen** Conceptualization, Methodology, Writing – Original Draft, Writing – Review & Editing, Supervision, Project administration, Funding acquisition.

## Statements and Declarations

Not applicable.

**Ethical considerations & Consent to participate** (also mentioned in methods)

## Funding

This research was partly funded by the European Union’s Horizon 2020 research and innovation program “ERA-NET rare disease research implementing IRDiRC objectives - N° 643578” via the Dutch research funding agency ZON-MW, through the E-Rare Joint Transnational Call JTC 2018 “Translational Research Projects on Rare Diseases” (ReCognitION project: Recognition and validation of druggable targets from the response to Cognitive Behavior Therapy in Myotonic Dystrophy type 1 patients from integrated -omics networks). This study was also partially funded by the European Union Seventh Framework Program, under grant agreement no. 305697 (the Observational Prolonged Trial In Myotonic dystrophy type 1 to Improve Quality of Life Standards, a Target Identification Collaboration [OPTIMISTIC] project) and by a Dutch Research Council (NWO) grant to the Netherlands X-omics Initiative (project 184.034.019). ED is supported by a Chercheur-boursier Junior 1 salary award from the Fonds de recherche du Québec-santé (FRQS-311186). AR received funding from the German Society of Muscular Diseases (DGM). TR, AR and VD acknowledge the financial support of the Myositis-Netz. HL receives support from the Canadian Institutes of Health Research (CIHR) for Foundation Grant FDN-167281 (Precision Health for Neuromuscular Diseases), Transnational Team Grant ERT-174211 (ProDGNE) and Network Grant OR2-189333 (NMD4C), from the Canada Foundation for Innovation (CFI-JELF 38412), the Canada Research Chairs program (Canada Research Chair in Neuromuscular Genomics and Health, 950-232279), the European Commission (Grant # 101080249) and the Canada Research Coordinating Committee New Frontiers in Research Fund (NFRFG-2022-00033) for SIMPATHIC, and from the Government of Canada Canada First Research Excellence Fund (CFREF) for the Brain-Heart Interconnectome (CFREF-2022-00007).

**Data availability** (also mentioned in methods)

