## Supplemental Materials for "Large-scale Proteomics Profiling of Peripheral Blood of DM1 patients identifies biomarkers for disease severity and functional capacity"

### Appendix

#### Methods

##### Mass spectrometry-based protein quantification of the OPTIMSTIC samples

*Proteomics sample preparation*
3 µl of each human serum sample was added to 40 µl lysis buffer containing 5% sodium dodecyl sulfate (SDS) and 50 mM triethylammonium bicarbonate (TEAB), pH 8.5. Proteins were reduced by addition of 15 mM dithiothreitol and incubation for 30 minutes at 55˚C and then alkylated by addition of 30 mM iodoacetamide and incubation for 15 minutes at RT in the dark. Phosphoric acid was added to a final concentration of 1.2% and subsequently samples were diluted 7-fold with binding buffer containing 90% methanol in 100 mM TEAB, pH 7.55. The samples were loaded on the 96-well S-Trap™ plate (Protifi), placed on top of a deepwell plate, and centrifuged for 2 min at 1,500 x g at RT. After protein binding, the S-trap™ plate was washed three times by adding 200 µl binding buffer and centrifugation for 2 min at 1,500 x g at room temperature. A new deepwell receiver plate was placed below the 96-well S-Trap™ plate and 50 mM TEAB containing trypsin (1/100, w/w) was added for digestion overnight at 37°C. Using centrifugation for 2 min at 1,500 x g, peptides were eluted in three times, first with 80 µl 50 mM TEAB, then with 80 µl 0.2% formic acid (FA) in water and finally with 80 µl 0.2% FA in water/acetonitrile (ACN) (50/50, v/v). Eluted peptides were dried completely by vacuum centrifugation. Dried peptides were redissolved in 100 µl 0.1% FA, the peptide concentration was determined on a Lunatic spectrophotometer (Unchained Labs) and was adjusted to 0.015 µg/µl with 0.1% FA[1]. iRT peptides (Biognosys, P/N Ki-3002-1) were added to all samples according to the manufacturer's instructions. Then, 300 ng of each sample was loaded on Evotips (Evosep, P/N EV2003) according to the manufacturer's instructions with a substitution of the wash step after sample loading by two wash steps with 80 µl 0.1% FA. All loaded Evotips were stored in 0.1% FA at 4°C until LC-MS/MS analysis was started.

*LC-MS/MS analysis*
Peptides from the human serum samples were run in data-independent acquisition (DIA) mode on an Evosep One LC-system (Evosep, Denmark) in-line connected to a Q Exactive HF mass spectrometer (Thermo). Peptides were analyzed with the 30 SPD method using the endurance Evosep column (15 cm x 150 µm I.D., 1.9µm beads, EV-1106, Evosep, Denmark) connected to a stainless steel emitter (30 µm inner diameter) (EV-1086, Evosep, Denmark). For elution of the peptides from the column 0.1% FA in LC-MS-grade water and 0.1% FA in ACN were used as mobile phases. Full-scan MS spectra ranging from 375-1500 m/z with an AGC target value of 5E6, a maximum fill time of 50 ms and a resolution at 200 m/z of 60,000 were followed by 30 quadrupole isolations with a precursor isolation width of 10 m/z for HCD fragmentation at an NCE of 30% after filling the trap at a target value of 3E6 for maximum injection time of 45 ms. MS2 spectra were acquired at a resolution of 15,000 at 200 m/z in the orbitrap analyser without multiplexing. The isolation intervals ranging from 400 – 900 m/z, with an overlap of 5 m/z were created with the Skyline software tool. The polydimethylcyclosiloxane background ion at 445.120028 Da was used for internal calibration (lock mass) and QCloud was used to control instrument longitudinal performance during the project[2].

*Proteomic data analysis*
A total of six gas-phase fractionated (GPF) samples, characterized by narrow-window pooling, were subjected to analysis utilizing the DIA-NN algorithm (version 1.8.1)[3]. This analysis employed a library-free mode, referencing the UniProt FASTA database, encompassing 20,783 sequences. Distinct

precursor mass ranges, spanning from 400 to 1000 m/z, were designated for each GPF fraction. The following parameters were used: enzyme specificity restricted to the C-terminal residues of arginine and lysine, fragment m/z constraints set between 200 and 1800, and one missed cleavage. Cysteine carbamidomethylation was set as a fixed modification. Only results with a false discovery rate (FDR) below 1% were kept.  Subsequent to the GPF analysis, the identification spectra were combined into a spectral library. This assembled library served as search space for the wide-window DIA samples using the identical DIA-NN version. The GPF-established parameters were preserved, with the exception of the precursor mass range. Furthermore, the "match between runs" (MBR) functionality was activated during this phase of analysis.

**Mass spectrometry-based protein quantification of the Canadian and German cohort samples**

*Proteomics sample preparation*

Human serum samples were digested as described previously with minor modifications[4]. 2 µl serum was diluted 7.5-fold in 50 mM ammonium bicarbonate buffer in MS-grade water (ABC) of which 2 µl was taken for digestion (~20 µg total protein). Proteins were denatured by diluting the samples 1:1 with 8M urea/10 mM Tris pH 8.0, after which proteins were reduced by addition of 1 µl 10 mM DTT for 30 min at RT. Reduced cysteines were alkylated by addition of 1 µl 50 mM 2-chloroacetamide (CAA) and incubation in the dark for 30 min at RT. Samples were diluted with 3 volumes of 50 mM ABC after which 1 µg trypsin was added for overnight digestion at 37 °C. To quench the reaction, samples were diluted 1:1 with 2 % trifluoroacetic acid (TFA) to an end volume of 132 µl. 1 µl of each sample (equivalent of ~150 ng total protein) was loaded on Evotips (Evosep), together with iRT standards (Biognosys) according to the manufacturer's instructions. Directly after loading the Evotips, LC-MS/MS analysis was started.

*LC-MS/MS analysis*

Samples were analyzed in duplicate. Peptides were separated on an Evosep One liquid chromatography (LC) system using C18 Evotips (Evosep, Odense, Denmark). Peptide separation was performed on a 15 cm C18 column (Evosep Performance, ReproSil-Pur C18, 150 µm I.D., 1.5 µm particle size) and eluted with a linear gradient of buffer A (HPLC grade water with 0.1 % formic acid) and buffer B (acetonitrile with 0.1 % formic acid) according to the Evosep 30SPD method. The eluted peptides were analyzed on a timsTOF Pro 2 mass spectrometer (MS) (Bruker Daltonics, Bremen, Germany) operated in the standard data independent acquisition Parallel Accumulation-SErial Fragmentation (diaPASEF) mode for long gradients.  In DIA mode, MS/MS scans were recorded within a m/z range of 400–1,200 and an ion mobility range of 0.6–1.6 1/K0.

*Proteomic data analysis*

Protein identification and quantification analysis were done with Bruker Proteoscape (BPS, version 2025b, Bruker Daltonics) using TIMS DIA-NN[5]. Mass spectra were streamed via the BPS plugin directly from the timsTOF's acquisition control software (timsControl) to the BPS workstation via a dedicated LAN connection and pre-processed into a binary file for consumption by TIMS DIA-NN. A spectral library consisting of 11641 precursors, including peptide modification such as oxidation of methionine was re-annotated against the canonical Uniprot human protein database (downloaded on 16-8-2024) plus sequences of known contaminants such as keratin and porcine trypsin. 20 ppm precursor tolerance and 15 ppm fragment ion tolerance was used and maxLFQ intensities were calculated. Multiple measurements were assembled and match-between-runs performed to fill-in missing values with an outlier frequency of 0.2, protein grouping was performed on the protein name level, no normalization was performed.

#### Results / Supplemental figures

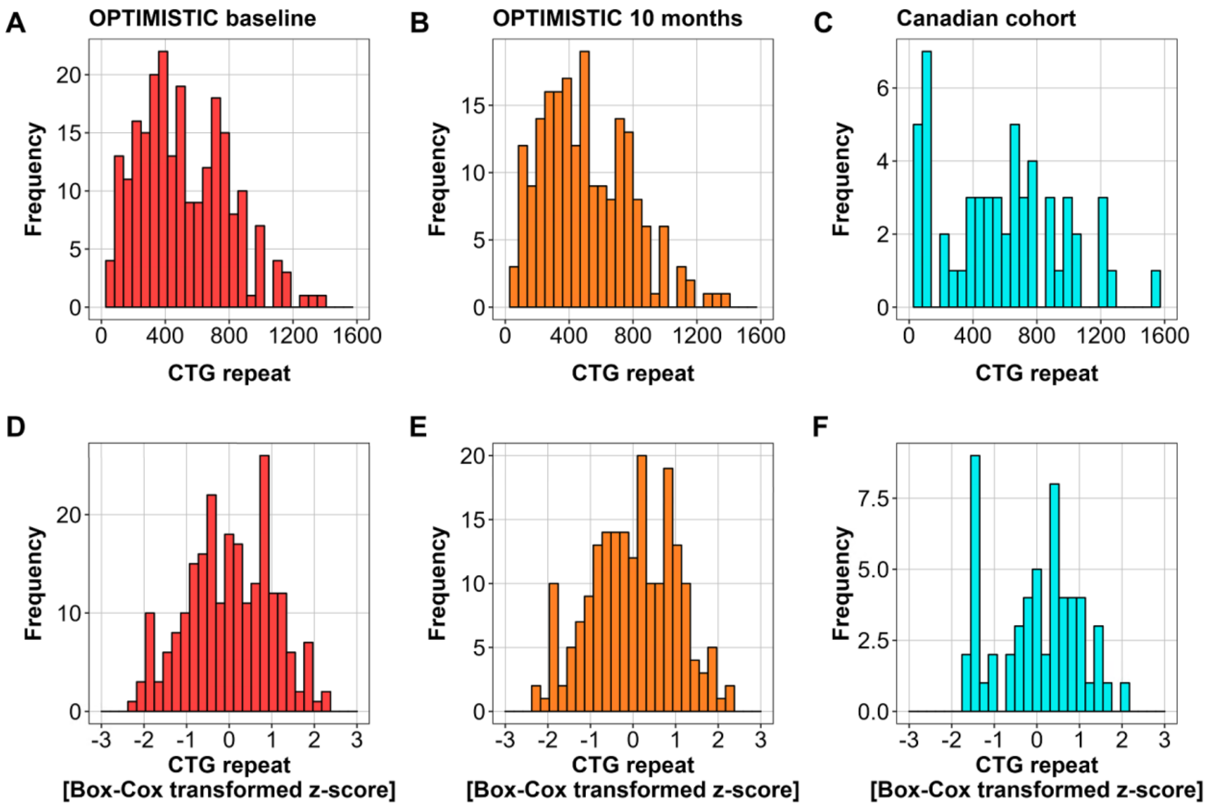


**SFigure 1*:* Differences in CTG repeat distributions across the different patient cohorts**.
Panels A, B, C show the distribution of the original CTG-repeats, while panels D, E, F show the distribution of the CTG-repeats after Box-Cox transformation and z-score standardization.

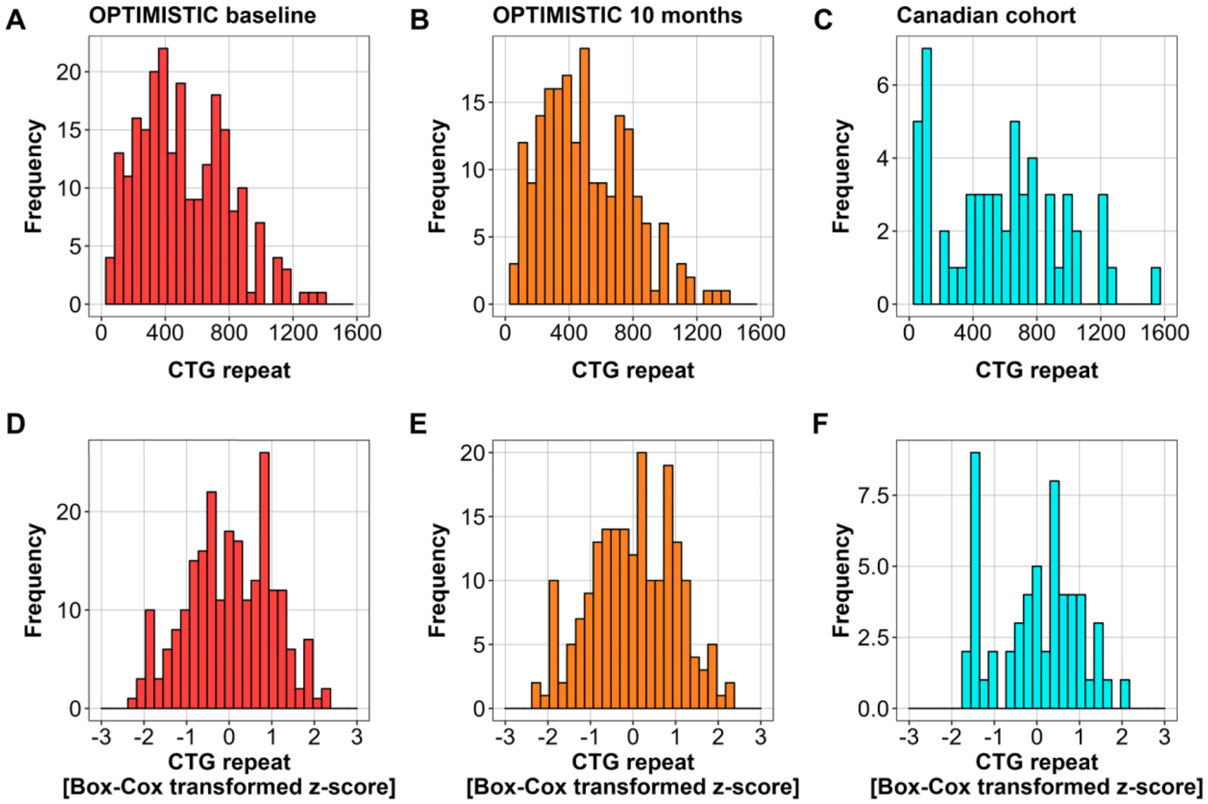


**SFigure 2*:* Differences in 6MWT distributions across the different patient cohorts**.
Panels A, B, C show the distribution of the original 6MWT scores, while panels D, E, F show the distribution of the 6MWT scores after Box-Cox transformation and z-score standardization.

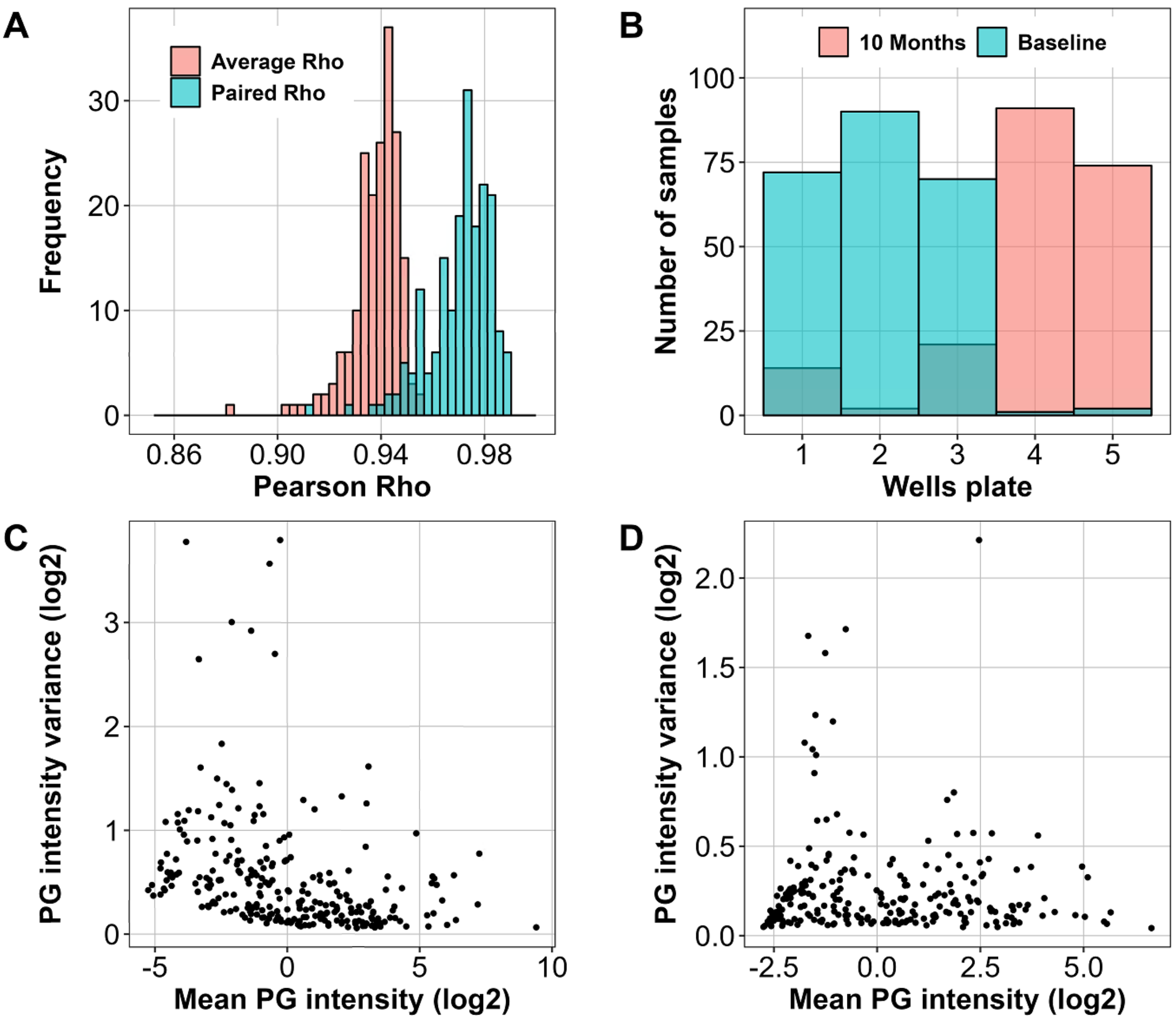


**SFigure 3: Quality analyses of protein quantifications.**

For the 189 paired OPTIMISTIC samples (same patient, two timepoints), the Pearson correlation was calculated for each protein group between the baseline and the matched 10-month sample (mint blue) as well as for each protein group between the baseline and all other 10-month samples (average per protein group in coral, panel A). Panel B shows the distribution of the OPTIMISTIC serum samples across the five well plates before the Mass-spectrometry workflow. Panels C and D show the variance in protein group (PG) intensities plotted against the mean protein group intensities for respectively the OPTIMISTIC and external validation dataset after filtering, log2 transformation and median centring.

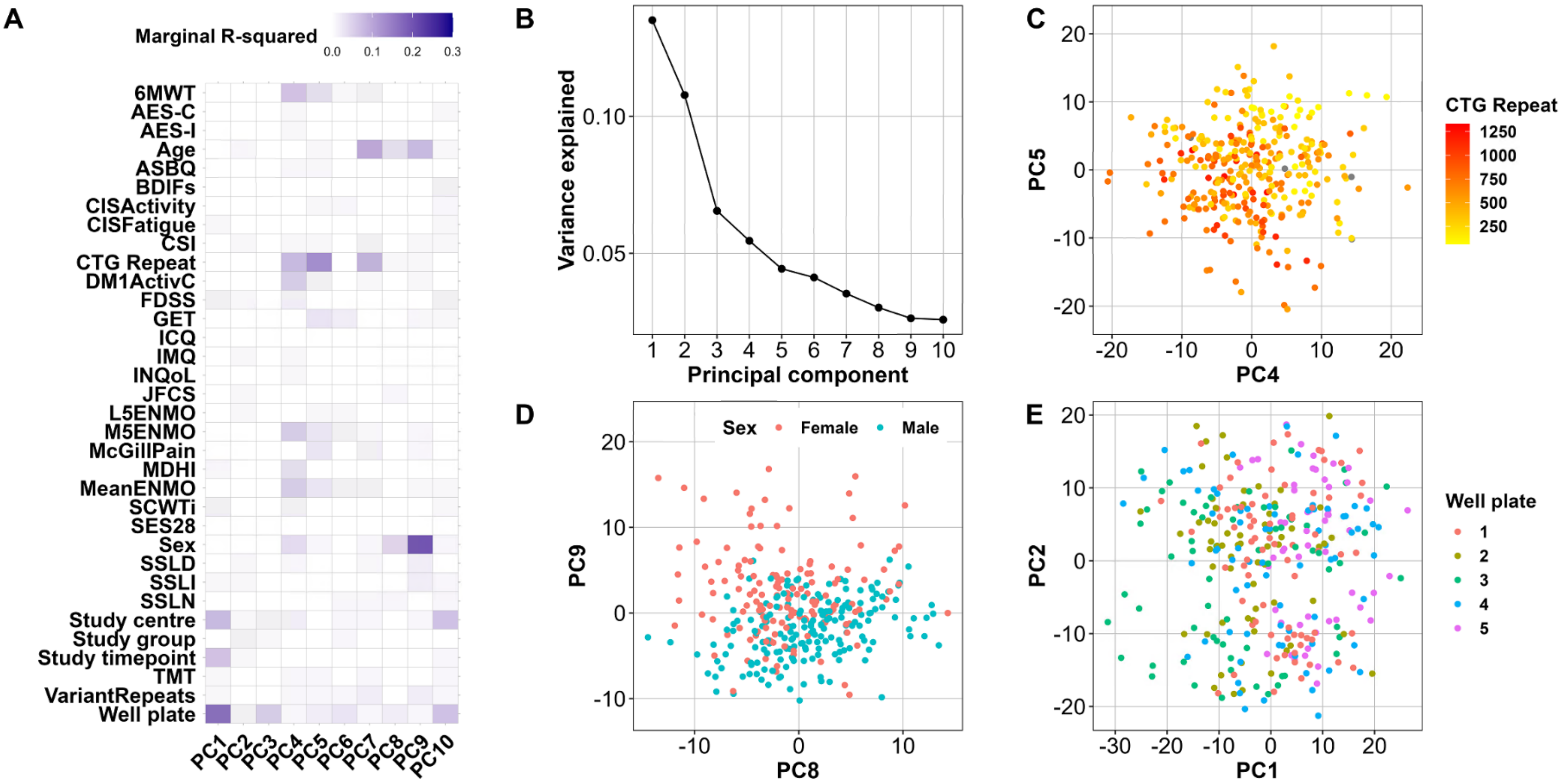


**SFigure 4: Drivers of peptide variance in OPTIMISTIC serum samples.**

A principal component analysis has been implemented on a filtered complete subset of the OPTIMISTIC peptide samples. Subsequently, for the first ten principal components (PC) a mixed effect model was fitted with various phenotype measures as predictor and PatientID as random effect (model 1). Associated marginal R-squared values are shown in panel A. Panel B shows the variance that is explained by the first 10 PC (Scree plot). The association of the CTG-repeat with PC4 and PC5, of the biological sex with PC8 and PC9 and of the well plate with PC1 and PC2 are respectively shown in panels C, D and E.

*Abbreviations: See Table 1 for outcome measures; GET Graded Exercise Therapy.*

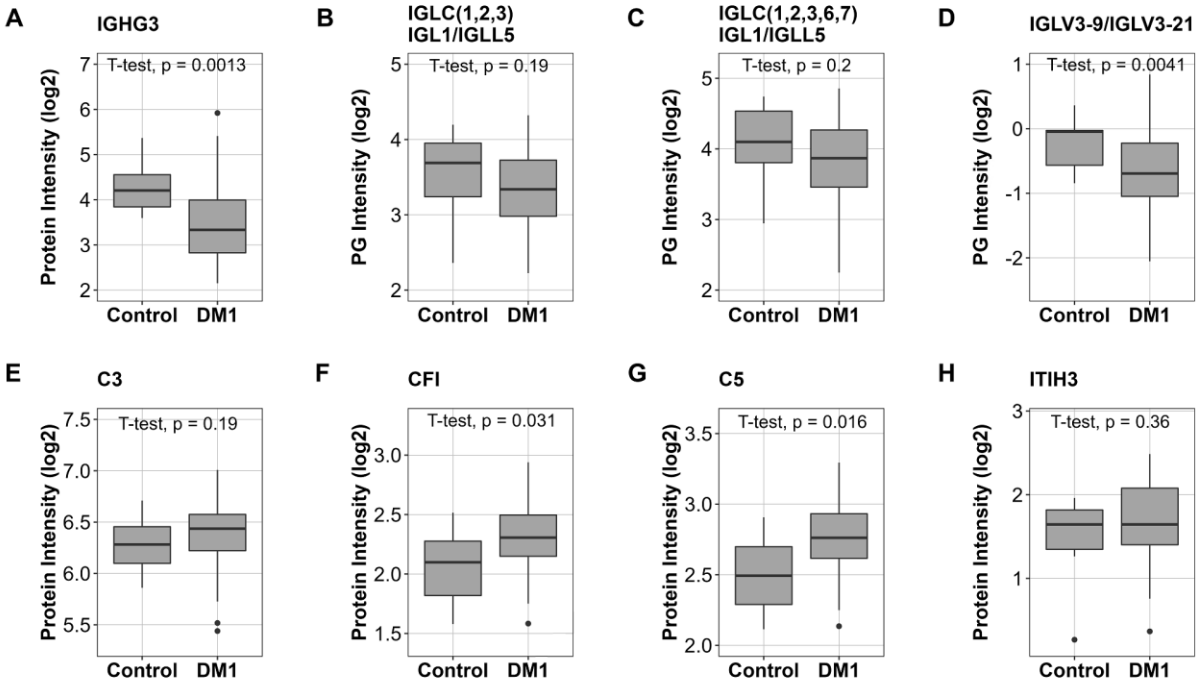


**SFigure 5: Protein group expression of top identified biomarkers in DM1 patients versus healthy controls**

Based on the full external validation dataset, consisting of MS-based serum protein group quantification of 56 Canadian DM1 patients, 13 German DM1 patients and 10 healthy controls, comparative plots were generated for the top (highest absolute Pearson Rho) identified immunoglobulin biomarkers associated with the CTG-repeat (panels A-D), for the top identified complement biomarkers associated with the 6MWT score (panels E-G) and for the ITIH3 6MWT association (panel H). Unpaired t-tests were implemented to study the significance of the different expression levels.

*Abbreviations: PG Protein group; IGHG3 Immunoglobulin heavy constant gamma 3; IGLC(1, 2, 3, 6, 7) Immunoglobulin lambda constant 1, 2, 3, 6, 7; IGL1 Immunoglobulin lambda-1 light chain; IGLL5 Immunoglobulin lambda-like polypeptide 5; IGLV3-9/3-21 Immunoglobulin lambda variable 3-9/3-21; C3 Complement C3; CFI Complement factor I; C5 Complement C5; ITIH3 Inter-alpha-trypsin inhibitor heavy chain H3.*

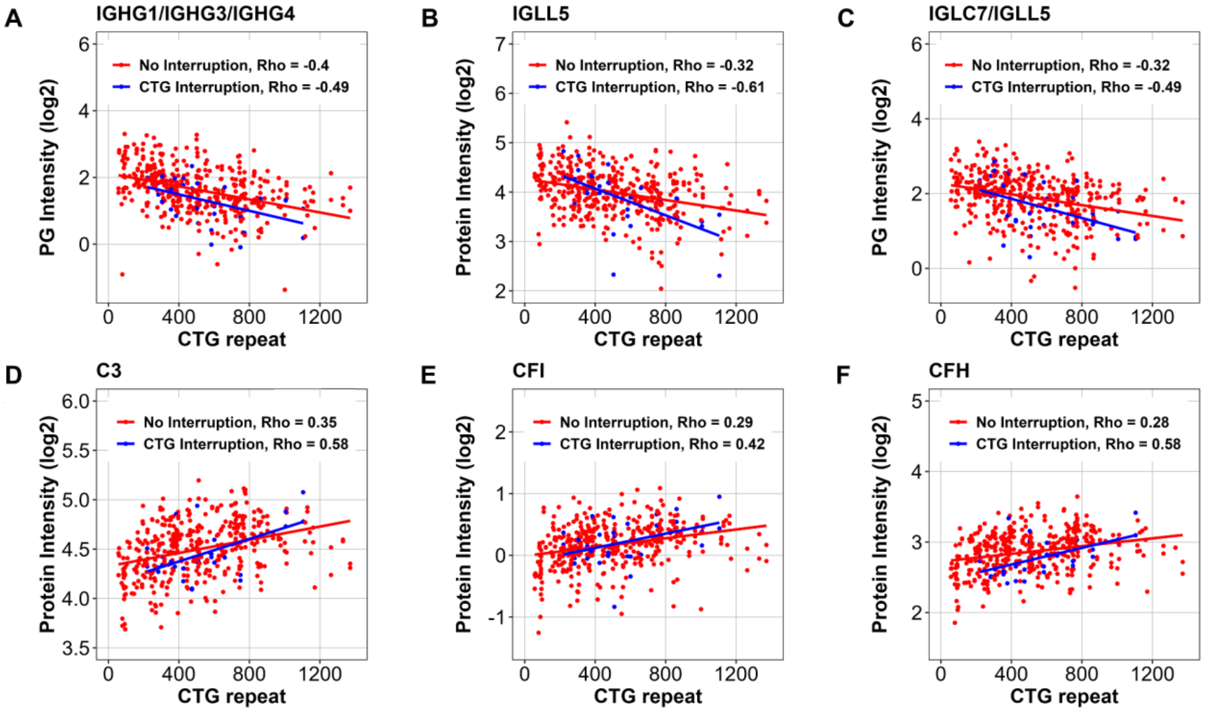


**SFigure 6*:* Effect of the CTG-repeat interruptions on the top CTG-repeat length associated protein groups.**

Information on the CTG-repeat interruptions was present for 436 of the OPTMISTIC serum samples, with 36 patients having a known repeat interruption (variant repeat). Depicted here are the three protein groups with the highest absolute Pearson rho values among the significant and validated CTG-repeat associations with gamma globulins (A-C) and the complement system (D-F), colour coded for whether or not a CTG-repeat interruption was present (red = no interruption, blue = interruption).

*Abbreviations: PG Protein group; IGHG1/3/4 Immunoglobulin heavy constant gamma 1/3/4; IGLL5 Immunoglobulin lambda-like polypeptide 5; IGLC7 Immunoglobulin lambda constant 7; C3 Complement C3; CFI Complement factor I; CFH Complement factor H.*

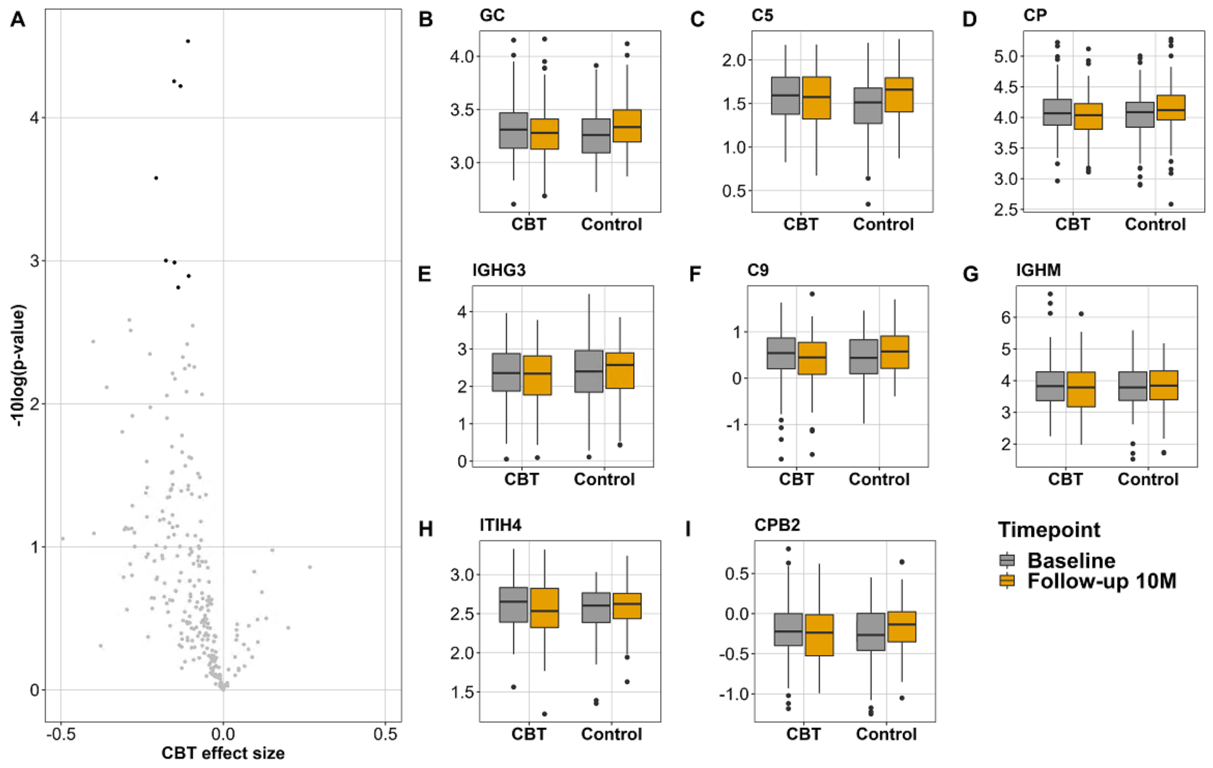


A

**SFigure 7: CBT effects on the serum proteome of DM1 patients**

For each identified protein group in the OPTIMISTIC serum samples, a mixed effect model (model 4) was fitted with the interaction effect of study timepoint (baseline / 10 months) with study intervention group (CBT/Control) as predictor, the covariate sex as fixed and PatientID and well plate as random effects. Panel A shows the volcano plot of the significance (-log10 of nominal p-values) and the interaction effect (CBT effect size), where significant associations (FDR < 0.05) are colour coded in black. For the eight significant associations (FDR < 0.05) boxplots were generated to illustrate the differences in protein expression per study timepoint and intervention group (panels B-F).

*Abbreviations: GC Vitamin D-binding protein; C5 Complement C5; CP Ceruloplasmin; IGHG3 Immunoglobulin heavy constant gamma 3; C9 Complement C9; IGHM Immunoglobulin heavy constant mu; ITIH4 Inter-alpha-trypsin inhibitor heavy chain H4; CPB2 Carboxypeptidase B2.*

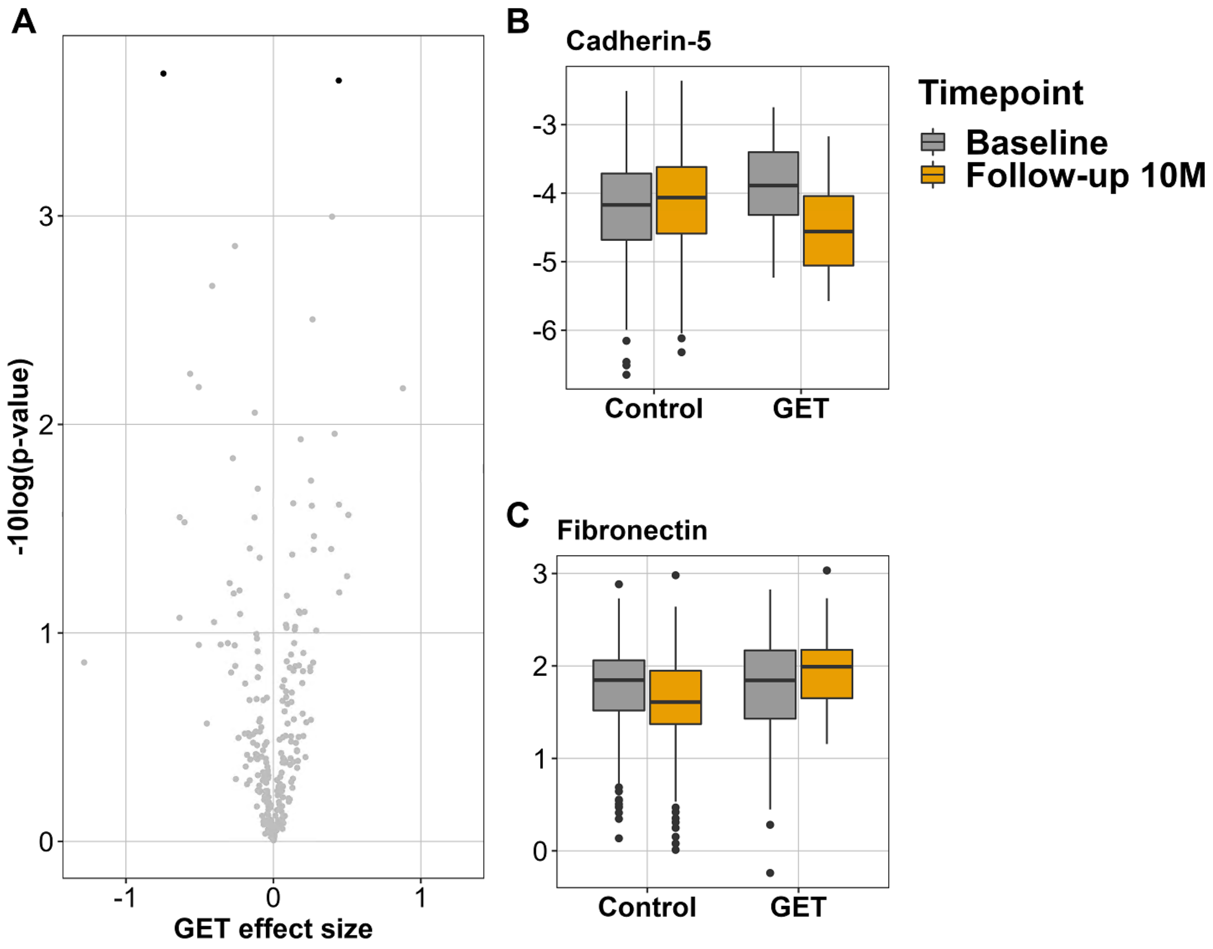


**SFigure 8: GET effects on the serum proteome of DM1 patients**

For each identified protein group in the OPTIMISTIC serum samples, a mixed effect model (model 5) was fitted with the interaction effect of study timepoint (baseline / 10 months) with study intervention group (Graded Exercise Therapy (GET/Control)) as predictor, the covariate sex as fixed and PatientID and well plate as random effects. Panel A shows the volcano plot of the significance (-log10 of nominal p-values) and the interaction effect (GET effect size), where significant associations (FDR < 0.05) are colour coded in black. For the two significant associations (FDR < 0.05) boxplots were generated to illustrate the differences in protein expression per study timepoint and intervention group (panels B-C).

5. Demichev V, Szyrwiel L, Yu F, et al. dia-PASEF data analysis using FragPipe and DIA-NN for

deep proteomics of low sample amounts. *Nat Commun 2022 131* 2022; 13: 1–8.
